# Multi-ancestry Polygenic Mechanisms of Type 2 Diabetes Elucidate Disease Processes and Clinical Heterogeneity

**DOI:** 10.1101/2023.09.28.23296294

**Authors:** Kirk Smith, Aaron J. Deutsch, Carolyn McGrail, Hyunkyung Kim, Sarah Hsu, Ravi Mandla, Philip H. Schroeder, Kenneth E. Westerman, Lukasz Szczerbinski, Timothy D. Majarian, Varinderpal Kaur, Alice Williamson, Melina Claussnitzer, Jose C. Florez, Alisa K. Manning, Josep M. Mercader, Kyle J. Gaulton, Miriam S. Udler

**Author notes:** contributed equally (Kirk Smith, Aaron J. Deutsch). Present address: TDM: Vertex Pharmaceuticals, Boston, MA, USA. Corresponding author: Miriam S. Udler.

## Abstract

We identified genetic subtypes of type 2 diabetes (T2D) by analyzing genetic data from diverse groups, including non-European populations. We implemented soft clustering with 650 T2D-associated genetic variants, capturing known and novel T2D subtypes with distinct cardiometabolic trait associations. The twelve genetic clusters were distinctively enriched for single-cell regulatory regions. Polygenic scores derived from the clusters differed in distribution between ancestry groups, including a significantly higher proportion of lipodystrophy-related polygenic risk in East Asian ancestry. T2D risk was equivalent at a BMI of 30 kg/m^2^ in the European subpopulation and 24.2 (22.9- 25.5) kg/m^2^ in the East Asian subpopulation; after adjusting for cluster-specific genetic risk, the equivalent BMI threshold increased to 28.5 (27.1-30.0) kg/m^2^ in the East Asian group, explaining about 75% of the difference in BMI thresholds. Thus, these multi-ancestry T2D genetic subtypes encompass a broader range of biological mechanisms and help explain ancestry-associated differences in T2D risk profiles.

## Introduction

The pathophysiology of type 2 diabetes (T2D) is influenced by multiple biological pathways, such as insulin resistance and beta cell dysfunction. Ongoing efforts have focused on advancing precision medicine in diabetes by identifying unique clinical trajectories or treatment approaches based on diabetes subtype^1^. Multiple strategies have been applied to identify T2D subtypes; some strategies rely on readily available clinical features and biomarkers, while other strategies incorporate additional information such as genomic data^2^.

Previously, we used genetic data and implemented a soft clustering approach using Bayesian non-negative matrix factorization (bNMF) to identify five T2D genetic subtypes^3^. Two subtypes were related to insulin deficiency (with elevated or decreased proinsulin levels), while three were related to insulin resistance, mediated by obesity, lipodystrophy, or abnormal liver/lipid metabolism. More recently, we developed a high-throughput pipeline to analyze a larger set of genome-wide association studies (GWAS) to increase our power to detect T2D subtypes^4^. This approach recapitulated our previous five clusters and also identified five additional clusters: another cluster related to beta cell dysfunction, a cluster with insulin resistance and very elevated insulin levels, and three clusters with abnormalities in alkaline phosphatase (ALP), sex hormone-binding globulin (SHBG), and lipoprotein A (LpA).

In this study, we expanded our analysis pipeline to investigate T2D genetic clusters using multi-ancestry cohorts. Previously, we focused on genetic data from individuals with European genetic ancestry; this was done due to limited data availability in other ancestry groups, as well as methodological limitations when analyzing genetic data from diverse populations. We hypothesized that incorporating individuals from diverse populations would increase the breadth of biological mechanisms captured and potentially explain ancestry-associated differences in T2D risk while avoiding the exacerbation of health disparities^5^. Thus, we leveraged the recent completion of additional multi-ancestry genetic studies^6–8^ to investigate T2D genetic clusters in diverse ancestral populations. Using an expanded set of cohorts, we replicated our previous T2D genetic clusters and identified new clusters related to decreased cholesterol levels, abnormal bilirubin metabolism, and abnormal lipid processing in adipose and hepatic tissues. We confirmed that common polygenic pathways contribute to T2D risk across multiple ancestral populations and are distinctively enriched for tissue-and single-cell regulatory regions. Additionally, we described associations of the genetic clusters with clinical phenotypes in two independent datasets, thereby characterizing the genetic subtypes. Finally, we demonstrated that the relative contribution of each genetic cluster differed according to ancestry group. This finding may help explain why certain individuals of a particular population genetic ancestry are more susceptible to T2D despite having a lower body mass index (BMI).

## Results

### Multi-ancestry clustering approach identifies twelve T2D genetic clusters

In our prior work, we developed a high-throughput pipeline to generate T2D clusters using T2D GWAS summary statistics^4^. Here, we expanded our set of input T2D GWAS to include those participants with non-European ancestry (Supplementary Table 1), and we updated our pipeline to account for varying allele frequencies across ancestry groups (Extended Data Fig. 1). We included a total of 37 T2D GWAS representing more than 1.4 million individuals across varied genetic ancestral backgrounds: African/African American, Admixed American, East Asian, European, South Asian, or multi-ancestry (Supplementary Table 1). After quality control and linkage disequilibrium (LD) pruning, we obtained a final set of 650 variants that had independent genome-wide significant associations with T2D (Supplementary Table 2). Next, we assembled a list of 165 GWAS of T2D-associated traits (Supplementary Table 3). After performing quality control and removing highly correlated traits, we assembled a final list of 110 traits. We then constructed an input matrix with our final set of 650 variants x 110 traits and applied our bNMF clustering algorithm.

The bNMF algorithm identified a total of twelve T2D genetic clusters (Fig. 1, Table 1, Supplementary Table 4). We identified three novel clusters, labeled as Lipodystrophy 2, Cholesterol, and Bilirubin. Compared to our prior work^4^, the multi-ancestry approach recaptured eight out of ten clusters: Beta Cell 1, Beta Cell 2, Proinsulin, Obesity, ALP Negative, Hyper Insulin, Lipodystrophy 1, and Liver-Lipid (Extended Data Fig. 2). The two remaining clusters from our prior work (SHBG and LpA) collapsed into a single cluster, denoted here as SHBG-LpA.

**Fig. 1.**
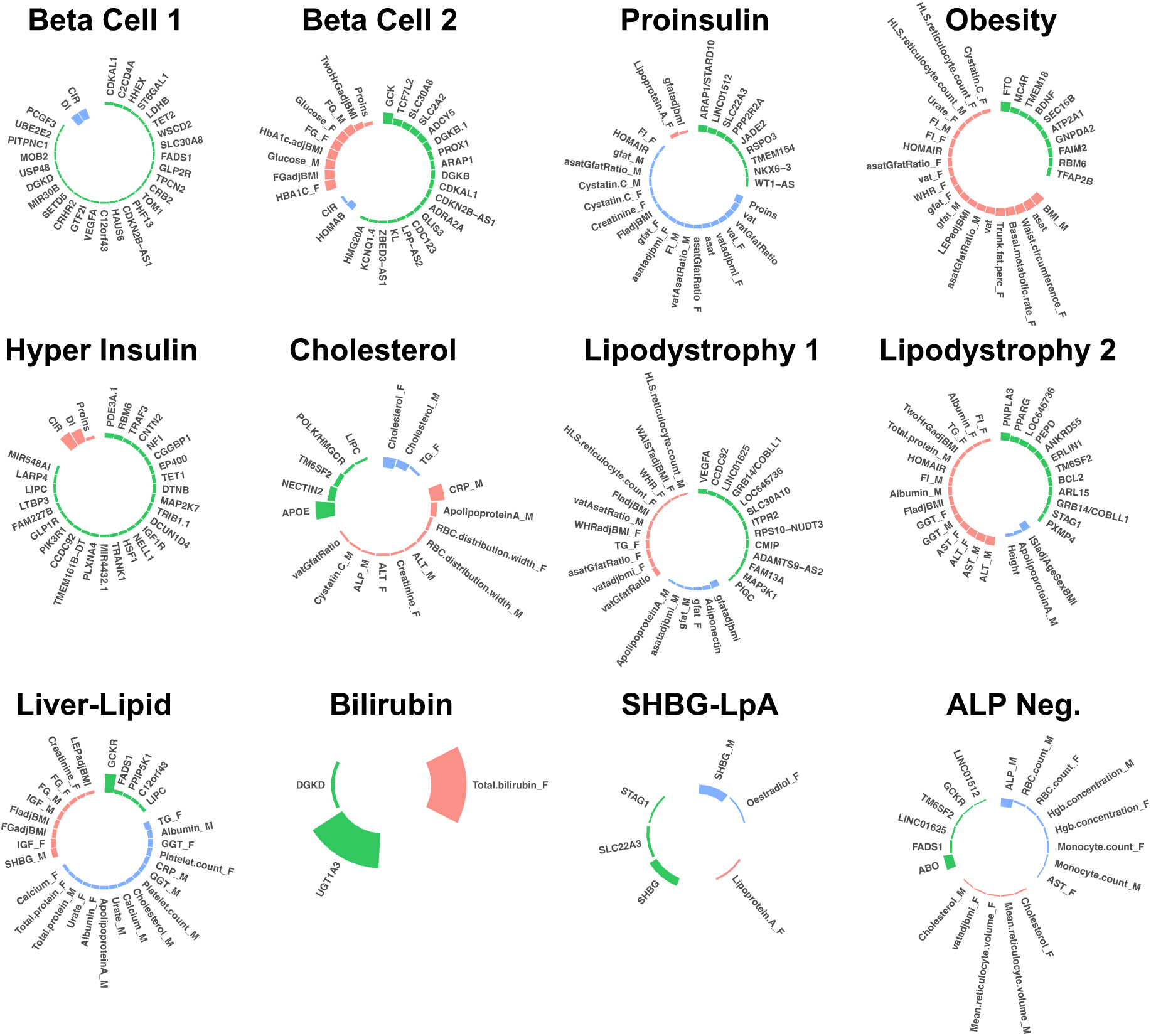
Key loci and traits of multi-ancestry T2D genetic clusters. Each plot displays the top-weighted loci and traits within each multi-ancestry T2D genetic cluster. The length of the bars corresponds to the cluster weight determined by the bNMF algorithm. Green bars represent genetic loci, red bars represent traits with increased values, and blue bars represent traits with decreased values within each cluster. Female-and male-specific traits are appended with “_F” and “_M”, respectively. A maximum of 30 elements (loci and traits) with the highest weights are displayed in each cluster.

**Table 1.** Overview of multi-ancestry T2D genetic clusters.

| Cluster (# Variants) | Expected physiologic al impact | Key top-weighted traits | Key top-weighted loci | Suspected mechanism | Note |
| --- | --- | --- | --- | --- | --- |
| **Beta Cell 1 (82)** | Insulin deficiency | CIR (-), DI (-) | *CDKAL1, C2CD4A, HHEX, ST6GAL1, LDHB, TET2* | Beta cell function, glucose homeostasis | Recaptures part of Beta Cell cluster from Udler et al 2018 [3] and Beta Cell 1 from Kim et al 2022 [4] |
| **Beta Cell 2 (40)** | Insulin deficiency | HbA1c female (+), FGadjBMI (+), Glucose male (+) | *GCK, TCF7L2, SLC30A8, SLC2A2, ADCY5, DGKB* | Beta cell function, insulin processing | Recaptures part of Beta Cell cluster from Udler et al 2018 [3] and Beta Cell 2 from Kim et al 2022 [4] |
| **Proinsulin (16)** | Insulin deficiency | PI (-), VAT (-) | *ARAP1/STARD10, VEGFA* | Insulin synthesis | Recaptures Proinsulin cluster from Udler et al 2018 [3] and Kim et al 2022 [4] |
| **Obesity (76)** | Insulin resistance | BMI male (+), ASAT (+), waist C female (+), Trunk fat % female (+) | *FTO, MC4R* | Obesity-mediated insulin resistance | Recaptures Obesity cluster from Udler et al 2018 [3] and Kim et al 2022 [4] |
| **Hyper Insulin (41)** | Insulin resistance | DI (+), CIR (+) | *PDE3A, RBM6, TRAF3, CNTN2* | Insulin secretion, inflammation | Recaptures Hyper Insulin cluster from Kim et al 2022 [4] |
| **Cholesterol (5)** | Unclear | CRP male (+), Cholesterol (-), Apolipoprotein A (+) | *APOE, NECTIN2, TM6SF2, POLK/HMGCR* | *HMGCR* expression | New cluster in this study |
| **Lipodystrophy 1 (47)** | Insulin resistance | GfatAdjBMI (-), vatGfatRatio (+), Adiponectin (-) | *VEGFA, CCFC92, CITED2, GRB14/COBLL1* | Fat distribution-mediated insulin resistance | Recaptures Lipodystrophy cluster from Udler et al 2018 [3] and Kim et al 2022 [4] |
| **Lipodystrophy 2 (29)** | Insulin resistance | ALT (+), ISladjAgeSexBMI (-), AST (+), GGT (+) | *PNPLA3, PARG, IRS1, PEPD, ANKRD55* | Hepatic steatosis | New cluster in this study; split from previous Lipodystrophy cluster |
| **Liver-Lipid (7)** | Insulin resistance | TG female (-), SHBG male (+), IGF female (+), Albumin male (-) | *GCKR, FADS1, PPIP5K1* | Liver/lipid metabolism | Recaptures Liver-Lipid cluster from Udler et al 2018 [3] and Kim et al 2022 [4] |
| **Bilirubin (2)** | Unclear | Bilirubin (+) | *UGT1A3* | Bilirubin metabolism | New cluster in this study |
| **SHBG-LpA (3)** | Unclear | SHBG male (-), Lp(a) female (+), oestradiol female (-) | *SHBG, SLC22A3, STAG1* | SHBG and Lp(a) metabolism | Merged from LpA and SHBG clusters from Kim et al 2022 [4] |
| **ALP Negative (6)** | Insulin resistance | ALP (-), RBC count (-), Hgb concentration (-) | *ABO, FADS1* | ALP activity levels | Recaptures ALP Neg cluster from Kim et al 2022 [4] |

The novel Lipodystrophy 2 cluster contained genetic determinants of lipid metabolism, liver dysfunction, and insulin resistance. The top-weighted traits included increased hepatic enzymes (aspartate aminotransferase [AST], alanine aminotransferase [ALT], γ-glutamyl transferase [GGT]), increased Homeostatic Model Assessment for Insulin Resistance (HOMA-IR), and decreased insulin sensitivity index. The top-weighted loci included *PNPLA3* and *PPARG*, which regulate the accumulation of fatty acids in the liver and adipose tissue^9,10^. Compared to our prior work, the previous single Lipodystrophy cluster appeared to split into two clusters (Extended Data Fig. 2), with traits related to body composition (such as increased visceral adipose tissue) remaining in the Lipodystrophy 1 cluster and traits related to hepatic function moving to the novel Lipodystrophy 2 cluster.

The novel Cholesterol cluster was associated with decreased LDL and total cholesterol levels. The top-weighted locus was *APOE*, which is directly involved in lipid metabolism^11^. Another top locus included the variant rs5744672, located on chromosome 5 near the *POLK* and *HMGCR* loci. *HMGCR* encodes HMG-CoA reductase, the enzyme responsible for the rate-limiting step of cholesterol synthesis and the target of statin medications. To support the hypothesis that the observed variation in cholesterol levels is mediated via *HMGCR*, we searched for expression quantitative trait loci (eQTL) in the Genotype-Tissue Expression Project (GTEx; gtexportal.org). We found a significant association between rs5744672 and *HMGCR* expression levels in skeletal muscle (normalized effect size = 0.14, *P* = 2.9 x 10^-6^), and there was strong LD between rs5744672 and the top eQTL variant for *HMGCR* (rs3846662, *r*^2^ = 0.91).

Finally, the novel Bilirubin cluster was associated with increased bilirubin levels. The cluster only included two variants, both located on chromosome 2 near the complex *UGT1A* locus, although the two variants were not in LD (*r*^2^ = 0.004). The *UGT1A* locus encodes multiple enzymes in the UDP-glucuronosyltransferase family, which are involved in excretion of bilirubin metabolites. We again looked for evidence to support the role of this genetic locus by searching for eQTLs in GTEx. We found that the top locus in the bilirubin cluster (rs887829) was significantly associated with gene expression of *UGT1A3* in the liver (normalized effect size = 0.46, *P* = 3.8 x 10^-14^); once again, there was strong LD between rs887829 and the top eQTL for *UGT1A3* (rs869283, *r*^2^ = 0.58).

### Common T2D genetic clusters are shared across individual ancestry groups

To determine whether the genetic clusters were specific to single populations or shared across populations, we repeated our clustering pipeline by focusing on each ancestry group individually. We found similar T2D genetic clusters within each ancestry group (Extended Data Fig. 3, Supplementary Tables 5-8). In summary, all groups included at least two clusters related to beta cell dysfunction and at least two clusters related to insulin resistance. The East Asian and European groups also captured clusters related to abnormal lipid metabolism in the liver and decreased ALP levels, which we also identified in our multi-ancestry clusters. We identified fewer clusters in the African and Admixed American groups, likely due to the fact that the GWAS sample sizes were smaller for these populations, leading to a smaller number of input genetic variants for the clustering algorithm. After confirming our findings in ancestry-specific analyses, we focused our remaining analyses on the multi-ancestry clusters, since the multi-ancestry clusters included all T2D GWAS from the ancestry-specific analyses and also included the trait GWAS with the largest sample sizes.

### Multi-ancestry clusters capture heterogeneous T2D genetic subtypes with distinct clinical associations

After creating the multi-ancestry T2D genetic clusters, we tested the association between individual clusters and specific clinical continuous traits or metabolic disease outcomes. To accomplish this, we implemented cluster-specific partitioned polygenic scores using individual-level data (pPS), or if not available, using published GWAS summary statistics (GWAS-pPS) (see Methods; Supplementary Table 9).

First, to characterize the clusters in terms of glycemic physiology, we assessed the associations of clusters with glycemic traits (Homeostatic Model Assessment of β-cell function [HOMA-B], HOMA-IR, proinsulin, and disposition index), using GWAS summary statistics included as inputs in the bNMF clustering algorithm (Fig. 2A, Supplementary Table 10). The Beta Cell 1, Beta Cell 2, and Proinsulin clusters were associated with decreased HOMA-B (β = -0.002 to -0.01, *P* < 1 x 10^-4^), suggesting a primary disease mechanism of insulin deficiency. The Obesity, Lipodystrophy 1, and Lipodystrophy 2 clusters were associated with increased HOMA-IR (β = 0.005 to 0.007, *P* < 1 x 10^-8^), suggesting a primary mechanism of insulin resistance. Two clusters (Liver-Lipid and ALP Negative) were associated with both decreased HOMA-B (β = -0.005 to -0.006, *P* < 4 x 10^-3^) and increased HOMA-IR (β = 0.005 to 0.007, *P* < 0.02), suggesting possible contributions from both mechanisms. The Hyper Insulin cluster was not significantly associated with HOMA-B or HOMA-IR levels, but it was characterized by a marked increase in the corrected insulin response (β = 0.05, *P* = 3.7 x 10^-17^). The final three clusters (Cholesterol, Bilirubin, and SHBG-LpA) were not clearly associated with mechanisms of insulin deficiency or resistance.

**Fig. 2.**
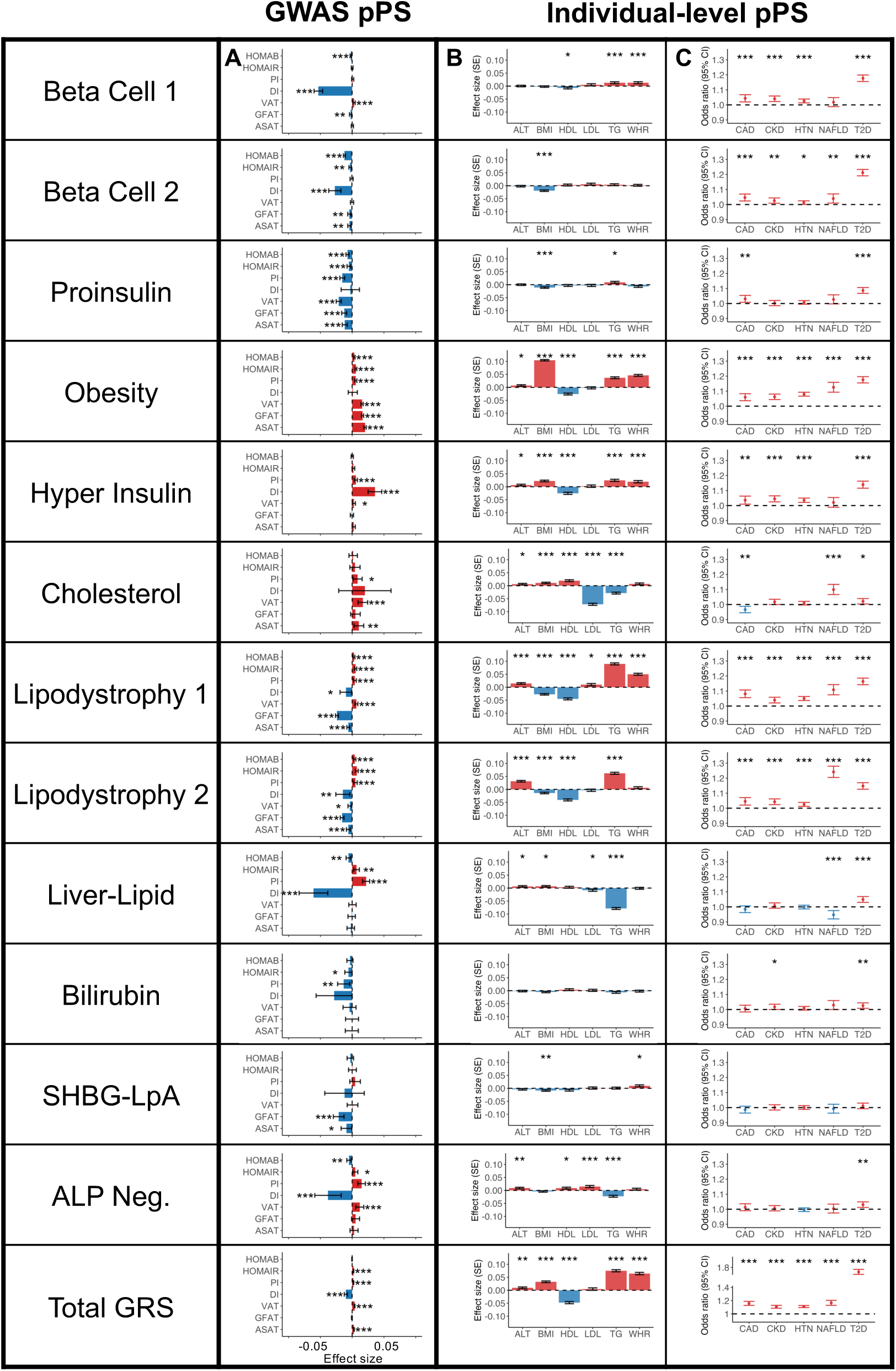
Multi-ancestry T2D genetic cluster associations with continuous traits and clinical phenotypes. (A) Each plot displays associations between selected multi-ancestry T2D genetic clusters and selected continuous outcomes, based on GWAS-partitioned pPS. The effect size indicates the beta coefficient from a meta-analysis of GWAS summary statistics. Error bars represent the 95% confidence interval. (B) Each plot displays cluster associations with selected continuous outcomes, based on individual-level pPS obtained from a meta-analysis of MGB Biobank and All of Us. Each outcome was normalized to a standard normal distribution. Effect sizes indicate the effect per one standard deviation increase in the pPS. Error bars represent the standard error from a linear regression model. (C) Each plot displays cluster-specific odds ratios of selected clinical phenotypes, based on individual-level pPS obtained from a meta-analysis of MGB Biobank and All of Us. Odd ratios are calculated per one standard deviation increase in the pPS. Error bars represent the 95% confidence interval. For all components, positive associations are colored in red, negative associations are colored in blue, and *P* values are indicated with asterisks (* *P* < 0.05, ** *P* < 0.01, *** *P* < 0.001).

Next, to assess how the T2D multi-ancestry clusters inform individual-level clinical differences, we calculated pPS in biobank participants. We performed our primary analysis in the All of Us cohort and replicated our findings in the Mass General Brigham (MGB) Biobank; here, we present our findings from a meta-analysis of over 100,000 participants (including over 14,000 with T2D) from the two cohorts (Supplementary Table 11).

First, in participants with and without T2D, we validated the relationship between cluster pPS and clinical measurements (Fig. 2B, Supplementary Tables 12-13). For example, we confirmed that the clusters had varied associations with lipid measurements: the Cholesterol pPS was associated with elevated HDL (β = 0.02 standard deviations [SD] of HDL per SD of pPS, *P* = 4.1 x 10^-8^) and decreased triglycerides (β = -0.03, *P* = 2.1 x 10^-15^), whereas the Lipodystrophy 1 pPS was associated with lower HDL (β = -0.04, *P* = 3.6 x 10^-33^) and higher triglycerides (β = 0.09, *P* = 1.2 x 10^-125^). These findings were largely consistent after adjusting for T2D status, or when analyzing the subset of individuals with T2D only (Supplementary Table 12).

Second, we tested the association between pPS and cardiometabolic phenotypes (chronic kidney disease [CKD], hypertension, coronary artery disease [CAD], non-alcoholic fatty liver disease [NAFLD], diabetic retinopathy, and diabetic neuropathy) (Fig. 2C, Supplementary Tables 14-15). The Lipodystrophy 2 cluster was associated with increased risk of NAFLD (OR = 1.24, *P* = 1.0 x 10^-44^), whereas the Liver-Lipid cluster was associated with decreased risk of NAFLD (OR = 0.95, *P* = 3.0 x 10^-4^); these findings remained after adjusting for T2D status (Fig. 2C, Supplementary Table 15). The Cholesterol cluster pPS was nominally associated with decreased risk of CAD (OR = 0.97, *P* = 2.3 x 10^-3^) in the individual-level data, and this negative association reached greater significance in the GWAS-pPS analysis (OR = 0.97, *P* = 2.8 x 10^-27^; Supplementary Table 10). The associations between other cluster pPS and cardiometabolic phenotypes were similar when we analyzed GWAS-pPS (Supplementary Table 10).

In the subset of individuals with T2D, the genetic clusters were also associated with significant clinical differences, as noted above (Supplementary Table 12). In this subgroup, we also analyzed the risk of two microvascular complications, diabetic retinopathy and diabetic neuropathy. No cluster was significantly associated with microvascular complications at the Bonferroni-adjusted threshold, although nominal associations were observed for the Lipodystrophy 1 cluster with diabetic retinopathy (OR = 1.09, *P* = 2.3 x 10^-3^) and the Obesity cluster with diabetic neuropathy (OR = 1.06, *P* = 8.5 x 10^-3^; Supplementary Table 15).

### Sex-stratified analyses of anthropometric and laboratory measures show distinct effects of multi-ancestry T2D genetic clusters in both sexes

Because lipid metabolism and adipose tissue distribution differ by sex^12,13^, we also performed sex-specific analyses for a subset of traits by performing separate regression models with female or male participants only. We found that the Cholesterol cluster was associated with decreased LDL levels in all participants, after adjusting for use of lipid-lowering medications; however, the effect size was more than twice as high in female participants (β_female_ = -0.09 SD of LDL per SD of Cholesterol pPS) compared to male participants (β_male_ = -0.04; *P*_interaction_ between sex and Cholesterol pPS = 4.0 x 10^-3^) (Supplementary Table 13). In addition, the Liver-Lipid cluster was significantly associated with decreased LDL levels in female participants (β_female_ = -0.02) but not in male participants (β_male_ = +0.007; *P*_interaction_ = 4.0 x 10^-5^).

Next, we analyzed individual-level measures of waist and hip circumference (only available in All of Us), as well as measures of subcutaneous adipose tissue (SAT) and visceral adipose tissue (VAT) (only available in a subset of approximately 9,000 MGB Biobank participants). In sex-specific analyses, the Lipodystrophy 1 pPS was associated with a lower SAT level in males (β_male_ = -0.07), but not in females (β_female_ = - 0.003; *P*_interaction_ = 0.01). However, the Lipodystrophy 1 pPS was associated with increased VAT/SAT ratio in both males (β_male_ = 0.07) and females (β_female_ = 0.08; *P*_interaction_ = 0.50) (Extended Data Fig. 4, Supplementary Table 13). Furthermore, the Lipodystrophy 1 pPS was associated with a significantly higher waist-hip ratio in both females and males, but the effect size was much lower in males (β_male_ = 0.02) compared to females (β_female_ = 0.08; *P*_interaction_ = 8.5 x 10^-20^) (Extended Data Fig. 5, Supplementary Table 13). Findings were similar after adjusting for BMI as a covariate.

Finally, we analyzed sex-specific associations between T2D genetic clusters and clinical outcomes (Supplementary Table 16). We found that the Beta Cell 1 pPS was significantly associated with hypertension in females (OR_female_ = 1.04), but not in males (OR_male_ = 1.02; *P*_interaction_ = 3.5 x 10^-3^). Conversely, the Beta Cell 2 pPS was significantly associated with CAD in males (OR_male_ = 1.06), but not in females (OR_female_ = 1.03; *P*_interaction_ = 4.7 x 10^-2^). The Obesity pPS was significantly associated with NAFLD in both males and females, but the effect size was considerably higher in females (OR_female_ = 1.17) compared to males (OR_male_ = 1.08; *P*_interaction_ = 3.1 x 10^-3^).

### Genetic variants contained in multi-ancestry T2D clusters are transcriptionally active in diabetes-related cell types

To further explore the biological mechanisms of the multi-ancestry T2D genetic clusters, we assessed for epigenomic evidence of transcriptional activity across a wide array of human tissue types. To perform this analysis, we used CATLAS, a repository of candidate gene regulatory elements found in regions with accessible chromatin in specific single-cell types^14^. As a secondary analysis, we also examined tissue-specific epigenomic data generated by the Roadmap Epigenomics Consortium^15^. We found cluster-specific enrichment of epigenomic annotations in biologically relevant tissues (Fig. 3, Supplementary Table 17). Interestingly, the Beta Cell 1 cluster was enriched for epigenomic annotations in a diverse range of cell types, while the Beta Cell 2 cluster was specifically characterized by pancreatic islet cell enrichment (False Discovery Rate [FDR] < 0.01) (Fig. 3A). These associations between the two Beta Cell clusters and epigenomic enrichment in pancreatic islets was also captured in the Roadmap analysis (Fig. 3B). Meanwhile, the Liver-Lipid cluster was enriched in fetal hepatoblasts (FDR < 0.01). Both the Lipodystrophy 1 and 2 clusters were enriched for regulatory activity in adipose tissue (FDR < 0.01). These findings confirm that the genetic variants captured by different clusters have distinct effects in specific tissue types, and these effects relate to suspected disease mechanisms.

**Fig. 3.**
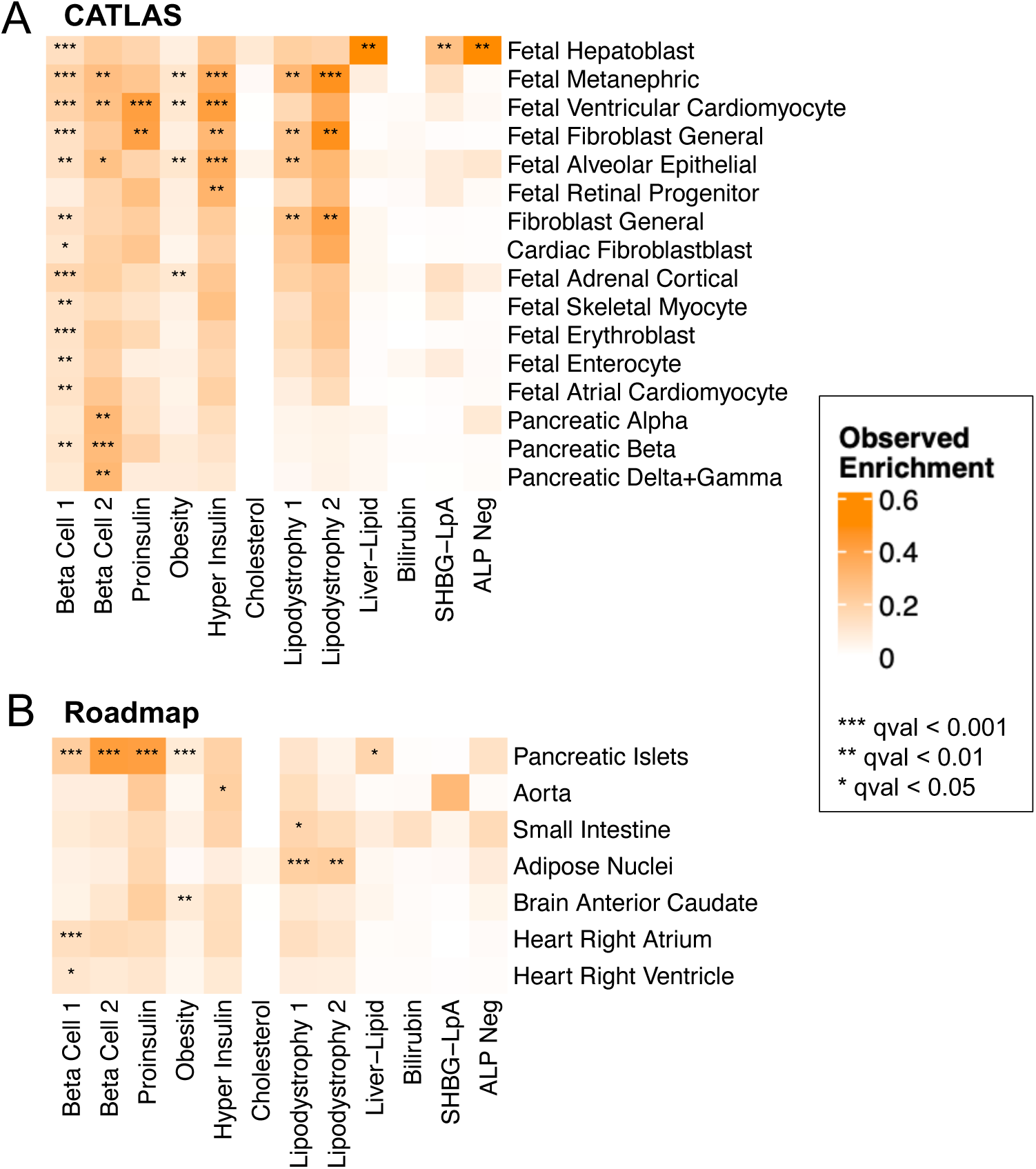
Enrichment for cell type specific enhancers in multi-ancestry type 2 diabetes clusters. Heatmaps display the significant cluster-specific enrichment of genomic annotations, represented by cumulative posterior probability, in (A) CATLAS single cell accessible chromatin data from 222 cell types and (B) Epigenomic Roadmap chromatin state calls from 28 cell types. *Q* values were corrected for false discovery rate (FDR). For both analyses, only cell types with at least one association of FDR < 0.1 are included in the figure, with additional data in Supplementary Table 17.

### Distribution of multi-ancestry T2D genetic clusters differs by genetic ancestry

Next, we assessed whether the multi-ancestry T2D genetic clusters had varying contributions to overall T2D genetic risk in individuals with different genetic ancestral backgrounds. We applied principal component analysis to classify individuals by genetic ancestry in both All of Us and MGB Biobank, using super-populations from the 1000 Genomes project: African (AFR), Admixed American (AMR), East Asian (EAS), European (EUR), South Asian (SAS), or other. After confirming that the distribution of T2D genetic clusters was similar in both cohorts, we then performed a meta-analysis of the two biobanks. For all individuals, we calculated pPS using the multi-ancestry T2D genetic clusters.

We found that the cluster-specific distribution of T2D genetic risk differed according to genetic ancestry. For example, the median Beta Cell 1 pPS was highest in the AFR ancestry group, whereas the median Lipodystrophy 1 and Lipodystrophy 2 pPS were highest in the EAS ancestry group (Fig. 4A; Extended Data Fig. 6). These differences led to varying proportions of individuals at the “extremes” of each genetic cluster. For instance, 10% of the EUR ancestry group had a Lipodystrophy 1 pPS above 1.11. However, 7.5% of the AFR group, 22.7% of the AMR group, 89.4% of the EAS group, and 28.2% of the SAS group had a Lipodystrophy 1 pPS above the same threshold (*P* < 10^-300^, one-way ANOVA). Furthermore, within each ancestry group, we found varied proportions of T2D genetic risk attributable to each cluster (Extended Data Fig. 7). This can be illustrated by the Lipodystrophy 1 cluster once again: 12.7% of the total T2D genetic risk across all clusters was present in the Lipodystrophy 1 cluster for the EAS ancestry group, compared to 8.1% for the AFR group, 9.6% for the AMR group, 9.0% for the EUR group, and 9.6% for the SAS group (*P* < 10^-300^, one-way ANOVA).

**Fig. 4.**
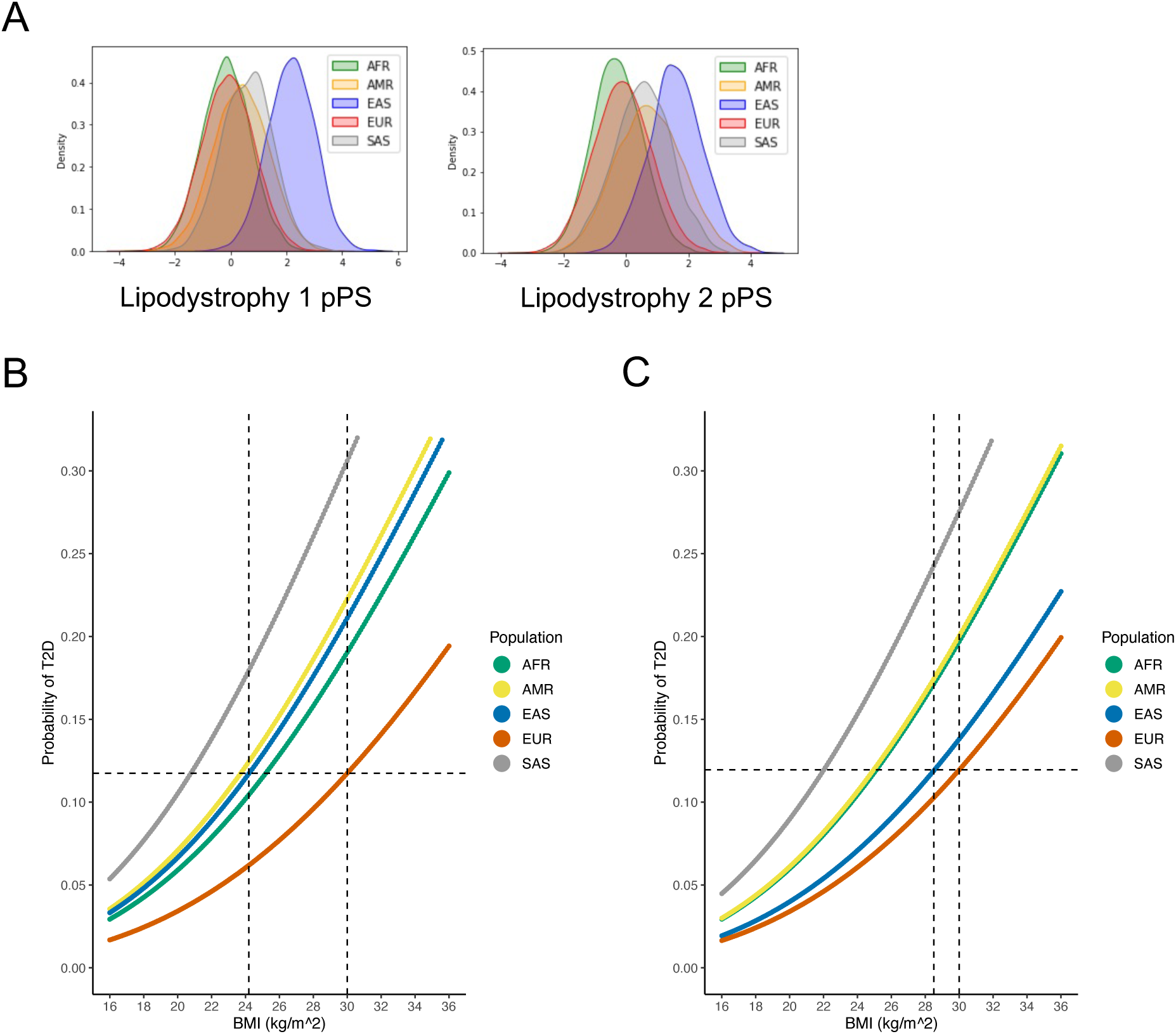
Ancestry-specific relationship between T2D genetic clusters, BMI, and T2D risk. (A) Ancestry-specific distribution of Lipodystrophy 1 and Lipodystrophy 2 pPS (normalized to a standard normal distribution). (B) Relationship between BMI and T2D risk (unadjusted), classified by genetic ancestry. The horizontal dashed line represents the T2D risk for participants with European genetic ancestry and a BMI of 30 kg/m^2^ (typically used to define obesity). The vertical dashed lines indicate the BMI thresholds needed to develop an equivalent risk of T2D in the European and East Asian ancestry groups. (C) Relationship between BMI and T2D risk, adjusted for Lipodystrophy 1 pPS and Lipodystrophy 2 pPS. All analyses were performed in a meta-analysis of MGB Biobank and All of Us.

### Differences in Lipodystrophy 1 and Lipodystrophy 2 genetic clusters explain increased risk of T2D in East Asian ancestry

We then investigated whether ancestry-specific variation in T2D genetic risk resulted in phenotypic differences between ancestry groups. We focused on the multi-ancestry Lipodystrophy 1 and Lipodystrophy 2 clusters, which were associated with decreased BMI but increased T2D risk (Fig. 2, Supplementary Table 12). After classifying individuals by genetic ancestry, we first analyzed the relationship between BMI and T2D risk in both All of Us and MGB Biobank separately; then, we performed a meta-analysis of both cohorts together.

Within each ancestry group, we calculated BMI thresholds with equivalent T2D risk. For example, at a BMI of 30 kg/m^2^ (typically used to define obesity), the risk of T2D within the EUR ancestry group was 11.7%. However, in order to achieve the same risk of T2D, the corresponding BMI cutoff was 25.2 kg/m^2^ (95% confidence interval 24.7-25.7) for the AFR group, 23.7 kg/m^2^ (23.1-24.3) for the AMR group, 24.2 kg/m^2^ (22.9-25.5) for the EAS group, and 20.8 kg/m^2^ (19.4-22.2) for the SAS group (Fig. 4B). After adjusting for the Lipodystrophy 1 and 2 pPS, to achieve the same risk of T2D as an individual with a BMI of 30 kg/m^2^ in the EUR ancestry group, the corresponding BMI cutoff was 25.1 kg/m^2^ (24.6-25.6) in the AFR group, 25.0 kg/m^2^ (24.4-25.5) in the AMR group, 28.5 kg/m^2^ (27.1-30.0) in the EAS group, and 22.0 kg/m^2^ (20.6-23.4) in the SAS group (Fig. 4C). Thus, by accounting for cluster-specific pPS, the difference in T2D risk-equivalent BMI thresholds between the EAS and EUR ancestry groups decreased by about 75%.

Next, we confirmed that the difference in T2D risk-equivalent BMI thresholds was primarily driven by the Lipodystrophy 1 and 2 clusters. When we adjusted for pPS from all 12 clusters simultaneously, the T2D risk was equivalent at a BMI of 30 kg/m^2^ in the EUR ancestry group and 28.3 kg/m^2^ (26.8-29.8) in the EAS group, compared to 28.5 kg/m^2^ (27.1-30.0) when adjusting for only the Lipodystrophy 1 and 2 clusters.

Finally, we assessed whether differences in adipose tissue distribution could account for variation in T2D risk. We analyzed a subset of 9,000 MGB Biobank participants with available VAT and SAT measurements. In a multivariate logistic regression model accounting for all 12 cluster pPS and BMI, the Lipodystrophy 1 and 2 pPS were significantly associated with T2D risk (Lipodystrophy 1: β = 0.11, *P* = 3.2 x 10^-3^; Lipodystrophy 2: β = 0.12, *P* = 6.3 x 10^-4^). However, after additionally accounting for the VAT/SAT ratio as a covariate, the association between both clusters and T2D risk was attenuated (Lipodystrophy 1: β = 0.06, *P* = 8.9 x 10^-2^; Lipodystrophy 2: β = 0.11, *P* = 1.8 x 10^-3^), suggesting that the T2D risk conferred by these clusters is partially mediated by changes in the VAT/SAT ratio (even after controlling for BMI).

## Discussion

In this study, we assembled a diverse set of GWAS to analyze 650 independent T2D-associated variants, as well as their associations with 110 relevant traits, and we identified twelve potential T2D genetic subtypes. By including genetic variants from multiple ancestry groups, we validated and expanded on our prior T2D clustering work, which focused on European populations and included only 323 variants^3,4^. We confirmed the existence of eight previously identified T2D genetic clusters, and we found that the previously defined SHBG and Lipoprotein A clusters^4^ merged into a single cluster. A new cluster, which we denoted as Lipodystrophy 2, split from the previous Lipodystrophy cluster, and we identified novel clusters associated with cholesterol and bilirubin. These clusters were significantly enriched in regulatory genomic regions in both bulk tissue and single cell epigenomic datasets. Additionally, we characterized the clinical features of the genetic subtypes derived from these clusters, including sex-specific analyses as well as association results with NAFLD for the first time.

The novel Lipodystrophy 2 cluster included multiple variants related to adipocyte and hepatocyte function. For instance, the cluster included variants near *PNPLA3*^9^ and *PPARG*^10^, which regulate fatty acid metabolism in both adipocytes and hepatocytes, and epigenomic analysis demonstrated that the Lipodystrophy 2 cluster was enriched in adipocytes (Fig. 3B). These findings support the concept that the Lipodystrophy 2 cluster split from the prior Lipodystrophy cluster (Extended Data Fig. 2); indeed, the Lipodystrophy 1 and 2 clusters in our current analysis shared multiple phenotypic similarities, such as decreased BMI, decreased HDL, and increased triglycerides, as well as increased risk of CAD, CKD, hypertension, and NAFLD. However, the clusters differed in their effects on body composition; only the Lipodystrophy 1 cluster (and not the Lipodystrophy 2 cluster) was associated with increased VAT and increased waist-hip ratio (Extended Data Figs 4-5, Supplementary Table 13).

A second new cluster was the Cholesterol cluster, which is notable because it reflects the complex relationship between T2D, coronary heart disease, and LDL cholesterol. While multiple genetic loci confer increased risk for both T2D and coronary heart disease^17,18^, we identified for the first time a cluster of T2D genetic variants that is associated with decreased risk of coronary heart disease. The Cholesterol cluster included an eQTL for *HMGCR*, the target of statin medications, which decrease the risk of coronary heart disease via lower LDL cholesterol. Hence, this cluster suggests that individuals with decreased expression in *HMGCR* have an increased risk of T2D, consistent with the observation that statin medications are associated with an increased risk of T2D^19^.

Aside from *HMGCR*, our multi-ancestry T2D genetic clusters confirmed the role of multiple genetic variants encoding proteins that serve as drug targets. For example, the Lipodystrophy 1 and 2 clusters included rs17036160 near *PPARG*, the target of thiazolidinediones, which promote insulin sensitivity. Furthermore, the Hyper Insulin cluster included rs10305420 near *GLP1R*, the target of GLP1 receptor agonists, which potentiate insulin secretion. Finally, the Lipodystrophy 1 cluster included rs998584 near *VEGF*, the target of VEGF inhibitors, which are used to treat diabetic retinopathy. Although the link between lipodystrophy and diabetic retinopathy is unclear, we found that an increased Lipodystrophy 1 pPS was nominally associated with an increased risk of diabetic retinopathy, and case studies have reported the occurrence of diabetic retinopathy in patients with congenital^20^ or acquired^21^ lipodystrophy.

The biological significance of the novel Bilirubin cluster is unclear. The top locus included an eQTL for *UGT1A3*, which is directly involved in bilirubin metabolism. Although the cluster suggests a positive association between bilirubin levels and T2D risk, previous studies have demonstrated a negative association^22^. In addition, bile acid sequestrants may be used for treatment of T2D^23^; however, the link between bile acid sequestrants and serum bilirubin levels is uncertain.

By adopting a multi-ancestry clustering approach, we captured the effects of variants that achieved genome-wide significance with T2D in multi-ancestry studies, but not in existing European-based studies. For instance, the Hyper Insulin cluster included rs59646751 near *IGF1R*, which had a *P* value of 2.4 x 10^-4^ in the European cohort of the MVP-DIAMANTE T2D GWAS and 3.4 x 10^-9^ in the multi-ancestry cohort^7^. This variant was identified in a previous exome sequencing study of T2D, where carriers of the variant had higher levels of circulating IGF-1, indicating IGF-1 resistance^24^. The assignment of this variant to the Hyper Insulin cluster suggests that carriers also have increased insulin resistance, possibly due to a negative feedback loop resulting in increased pituitary secretion of growth hormone.

This study also demonstrated novel sex-specific effects of T2D genetic clusters. For example, the association between the Cholesterol pPS and decreased LDL levels was primarily driven by a larger effect size in females. Similarly, the association between the Lipodystrophy 1 pPS and waist-hip ratio had a larger effect size in females. This suggests that these genetic associations may be driven by sex-specific physiology, potentially mediated by hormones that differ in males and females.

We also demonstrated how our multi-ancestry clusters can explain the heterogeneity of T2D among different populations. It is well-documented that individuals from various self-identified non-White populations are more susceptible to T2D at a given BMI, compared to self-identified White individuals^25,26^, and many authors have suggested that different BMI thresholds should be used to define obesity in separate populations^27−29^. Some studies have suggested that genetic differences in adipose tissue distribution may explain the varied relationship between BMI and T2D risk in different populations^30^. In this study, after classifying individuals by genetic ancestry, we confirmed prior obeservations^25,26^ that individuals in the EUR ancestry group had the lowest risk of T2D at all BMI strata. Furthermore, we demonstrated that variation in the BMI-T2D relationship is at least partially explained by variation in the distribution of T2D genetic risk among specific clusters. For instance, the EAS ancestry group had the highest Lipodystrophy 1 pPS as well as the highest Lipodystrophy 2 pPS (Fig. 4A). These clusters are associated with decreased BMI as well as increased T2D risk, resulting in a greater proportion of individuals with T2D at lower BMI levels. After adjusting for the

Lipodystrophy 1 pPS and the Lipodystrophy 2 pPS, the difference in BMI-adjusted T2D risk between the EAS and EUR ancestry groups was reduced by about **75%**, demonstrating the significant impact of these genetic clusters in East Asian populations (Fig. 4B, C). These findings represent a potential step toward developing individualized, genetically informed BMI recommendations for counseling patients to help prevent or ameliorate the effects of metabolic diseases. Notably, however, the Lipodystrophy 1 and Lipodystrophy 2 pPS were not markedly elevated in the other groups (AFR, AMR, and SAS) compared to the EUR subpopulation, so adjusting for these scores did not substantially affect T2D risk estimates. Thus, further work is needed to investigate genetic factors affecting the BMI-T2D relationship in these populations.

In parallel to our current study, the T2D Global Genomics Initiative (T2DGGI) also investigated T2D genetic clusters in multi-ancestry GWAS^31^. The T2DGGI clustering approach used genetic variants from a single, large multi-ancestry GWAS. In contrast, the study presented here used genetic variants from multiple ancestry-specific and multi-ancestry GWAS, including GWAS for T2D as well as for T2D-associated traits. Furthermore, the T2DGGI approach used a hard clustering method (K means clustering), compared to the soft clustering method presented here. Nevertheless, both studies found similar T2D genetic clusters. An analysis of the cluster weights assigned to both sets of genetic variants demonstrated high degrees of similarity in several clusters, including the Beta Cell, Obesity and Lipodystrophy 1 clusters (Extended Data Fig. 8). Despite the similarities between the two sets of T2D clusters, each study also included certain clusters that were not captured by the other study. For instance, our study included the Hyper Insulin, Bilirubin, SHBG-LpA, and ALP Negative clusters, whereas the T2DGGI study included clusters labeled as Body fat, Metabolic syndrome, and Residual glycemic. These findings support that there are multiple possible approaches to stratifying genetic loci, and further downstream analyses will be necessary to determine the utility of these approaches.

Overall, we demonstrated that similar patterns of T2D genetic subtypes occur across multiple populations. Using a multi-ancestry approach, we identified novel clusters that help to elucidate the complex relationship between T2D, CAD, and NAFLD. We also demonstrated how genetic variation across ancestry groups can cause differences in body fat composition, thereby altering T2D risk. To advance the care of patients with diabetes, current and future studies may focus on precision medicine strategies to target specific biological mechanisms highlighted by the T2D genetic clusters.

## Methods

### Pipeline for input variant–trait association matrix for clustering

The pipeline’s data preprocessing steps are detailed in the flowchart shown in Extended Data Fig. 1. For the multi-ancestry clusters, GWAS-significant (*P* < 5 x 10^-8^) variants were extracted from a diverse set of T2D GWAS (Supplementary Table 1), including studies performed in European, East Asian, African, Admixed American, South Asian and mixed cohorts. After removing indels and variants found in the major histocompatibility complex (MHC) region, variants underwent five independent iterations of LD-pruning (LD *r*^2^ < 0.05, MAF < 0.001), one for each population’s reference panel. Variants were only retained if found to be independent in all five populations. If any of the pruned variants had high-missingness across the trait GWAS, was multi-allelic or was ambiguous (A/T, C/G), then it was replaced with a high-LD (*r*^2^ > 0.8) proxy variant. As a final check, the variants were queried in the largest multi-ancestry T2D GWAS and were removed if they had *P* > 0.05 or if there were discrepancies in the noted risk alleles. The final set of 650 T2D-associated variants are shown in Supplementary Table 2.

For the traits included in the clustering, we compiled an extensive list of 165 continuous phenotypes GWAS and allowed the pipeline to determine which were relevant to the T2D variants (Supplementary Table 3). We prioritized sex-specific and multi-ancestry GWAS; however, if those were not available for a specific trait, then European-based GWAS were used. Traits were filtered out if their median sample size was below 5,000 or if their minimum *P* value for the final variant set was not Bonferroni-significant (*P*_min_ > 0.05/650 variants). Finally, we remove highly correlated traits (*R* > 0.80), prioritizing traits by their maximum variant-trait association (Supplementary Table 4C). With this final set of variants and traits (650 variants x 110 traits), we generated a matrix of standardized and scaled z-scores, which had been aligned to the T2D risk-increasing alleles. To fill in any remaining missing variant-trait associations in this final matrix, we used z-scores from proxies (LD *r*^2^ > 0.5) where possible, and otherwise assigned the trait’s median value.

The ancestry-specific clusters were generated using the same general steps; however, the input T2D GWAS were limited to studies where the cohort matched the population of interest. For the African and Admixed American clusters, the T2D *P* value threshold was lowered to *P* < 5 x 10^-6^, to account for the less powerful GWAS. The variants were pruned in a single iteration, using the appropriate reference panel for each population. For the traits, ancestry-specific GWAS were prioritized, followed by multi-ancestry and European-based summary statistics (Supplementary Table 3).

### Statistical comparison of cluster overlap

To compare different versions of the T2D genetic clusters, we focused on the cluster weights assigned to the T2D-related traits. For each pair of clusters, we calculated the Pearson correlation coefficient (*R*) between each set of trait cluster weights. We compared the multi-ancestry clusters generated in this study to the T2D genetic clusters identified in our prior studies^3,4^. We also compared the ancestry-specific clusters to the multi-ancestry clusters and to the T2D genetic clusters from our prior study^3^.

To compare the multi-ancestry clusters and the T2D clusters generated by the T2DGGI study^31^, we focused on the genetic loci included in each cluster, since the clustering method used by the T2DGGI study did not assign traits to specific clusters or generate cluster weights. First, we matched genetic variants included in the T2DGGI clusters to a corresponding high-LD variant (*r*^2^ > 0.5) from our multi-ancestry clusters. By doing so, we were able to transfer our variant weights to the T2DGGI clusters. We then assessed the correlation between genetic variant weights across the T2DGGI and multi-ancestry clusters using the Wilcoxon rank-sum test.

### Calculation of partitioned polygenic scores

We created partitioned polygenic scores (pPS) by calculating a weighted sum of the genetic variants in each cluster. We used individual-level data when possible; when unavailable, we used GWAS summary statistics. To calculate GWAS-partitioned pPS, we extracted the genetic variants from summary statistics of GWAS for specific traits. We combined the variants using inverse-variance weighted fixed effects meta-analysis, weighting each variant according to its GWAS effect size. We chose GWAS for several key glycemic traits (such as disposition index, proinsulin, and fasting insulin) as well as for measures of adipose tissue distribution or cardiometabolic outcomes. In addition, we calculated individual-level pPS using genotype data from two external biobanks: the All of Us research program^32^ and the Mass General Brigham (MGB) Biobank^33^. For individual-level pPS, we weighted the genetic variants according to the cluster weights generated by the bNMF algorithm. We only included those variants with a weight above 0.7802, a threshold which was calculated to maximize the signal-to-noise ratio, as described in Kim *et al.*^4^

### Biobank Analyses

For individual-level data, we performed a meta-analysis of two datasets. Each dataset was independent of the GWAS cohorts used to generate the clusters. Informed consent was obtained from all participants in both datasets. We complied with all relevant ethical regulations when analyzing genetic data from human research participants.

#### All of Us

Analysis of the All of Us cohort^32^ was approved by an institutional Data Use and Registration Agreement between MGB and the All of Us Research Program. We used the All of Us Controlled Tier Dataset v6. Full details on the demographic distribution of the dataset are provided in Supplementary Table 11. Individuals were classified as having type 2 diabetes if they were identified by an algorithm from Northwestern University as part of the Electronic Medical Records and Genomics (eMERGE) consortium^34^, or if they self-identified as having type 2 diabetes on the All of Us participant survey [Szczerbinski et al., manuscript in preparation]. The eMERGE algorithm classifies individuals based on diagnosis codes, medication prescriptions, and laboratory values. All individuals who were not classified as having type 2 diabetes were labeled as controls; however, individuals were excluded from the control group if they were less than 30 years old, or if they ever had a hemoglobin A1c greater than or equal to 6.5%. Other phenotypes were defined as described in Supplementary Table 14.

#### MGB Biobank

We used clinical and genomic data from the MGB Biobank^33^, which was current as of October 2022. Analysis of the MGB Biobank was approved by the MGB IRB (study protocol 2016P001018). Full details on the demographic distribution of the dataset are provided in Supplementary Table 11. Type 2 diabetes was defined using a phenotype algorithm developed by the MGB Biobank, with a set positive predictive value of 0.95. Once again, individuals were excluded from the control group if they were less than 30 years old, or if they ever had a hemoglobin A1c greater than or equal to 6.5%. Other phenotypes were defined as described in Supplementary Table 14.

### Assessment of transcriptional activity

We analyzed transcriptional activity of genetic loci using two databases of epigenomic information. For our primary analysis, we used CATLAS, a resource that maps regions of accessible chromatin across the human genome at single-cell resolution^14^. CATLAS uses ATAC-Seq to identify over 1 million candidate cis-regulatory elements across more than 200 distinct human cell types (both adult and fetal cells). As a secondary analysis, we used information from the Roadmap Epigenomics Consortium, which includes maps of regulatory elements for over 100 tissue types at the bulk tissue level^15^. To assess for enrichment of epigenomic annotations, we first defined 99% credible sets for each locus. To do this, we calculated approximate Bayes factors (aBFs) for all variants within a 500 kb window that had *r*^2^ ≥ 0.1 with the index variant, as described previously^35^. We calculated a posterior probability for each variant by dividing the aBF by the sum of all aBFs in the credible set. Next, within each cluster, we overlapped credible set variants with cell type genomic annotations and calculated the cumulative posterior probability (cPPA) for each annotation. We used a permutation test to assess the significance for annotations in each cluster. For each cluster, we permuted locus and cell type annotations and recalculated the cPPA based on shuffled labels. After performing 10,000 permutations, we compared the observed cPPA to the permuted background using a one-tailed test to determine the significance of each annotation. We corrected for multiple tests and defined statistically significant enrichment at *q* value thresholds of 0.1 and 0.001.

### Determination of genetic ancestry

We performed principal component analysis to uncover population stratification in each dataset (MGB Biobank and All of Us). Measurements that capture genetic similarity (such as principal components) are preferred when performing genomics research. However, due to privacy restrictions, we were unable to combine genomic data from both datasets to generate a single set of principal components. Therefore, we used principal component data to apply population descriptor labels at the level of continental ancestry, acknowledging that these labels are imprecise. We used a random forest classifier model to assign participants in each biobank to one of six continental ancestry groups (African, Admixed American, East Asian, European, Middle Eastern, or South Asian), following the method of the Pan-UK Biobank^36^. For any given individual, if the probability of each ancestry group was less than 50%, then the individual’s genetic ancestry was left as “unclassified”. The total number of individuals in each genetic ancestry group is listed in Supplementary Table 11. We excluded any population with fewer than 500 individuals in a given dataset; therefore, the Middle Eastern ancestry group was excluded from downstream analyses.

### Individual-level cluster associations with clinical phenotypes

After generating individual-level pPS, we analyzed the association of the pPS with various clinical phenotypes, using linear regression (for continuous outcomes) or logistic regression (for binary outcomes). We analyzed all associations in a meta-analysis of both biobanks (MGB Biobank and All of Us), using a random effects model. Each regression model was adjusted for the following covariates: age, sex, and genetically inferred ancestry. Certain regression models were also adjusted for type 2 diabetes status and/or BMI, as noted. A subset of analyses was performed separately for female or male participants only; these analyses included age and genetically inferred ancestry as covariates. To assess the significance of sex-specific differences, we constructed a regression model with all participants and included an interaction term between sex and cluster pPS. Clinical measurements that were not normally distributed were log-transformed to obtain a normal distribution^37^; following previous studies, these measurements included BMI and triglycerides^4,38^.

For validation tests that confirmed known associations between cluster pPS and variables used in the clustering algorithm, we did not use multiple test correction to denote statistical significance. Of note, clinical phenotypes were not directly used in the clustering algorithm, but several traits that were included in the clustering (i.e. glucose, hemoglobin A1c, creatinine, cystatin C, systolic blood pressure, and diastolic blood pressure) can define T2D, CKD, and hypertension. In contrast, association tests with the remaining phenotypes (CAD, NAFLD, diabetic retinopathy, and diabetic neuropathy) revealed novel associations with cluster pPS. For these discovery tests, we defined statistical significance using a Bonferroni-adjusted threshold of 0.05/(*K* x *N*), where *K* represents the number of clusters tested and *N* represents the number of phenotypes tested. For individual-level testing, we excluded any binary outcome in which fewer than 500 participants met the outcome in either biobank; therefore, ischemic stroke was excluded from downstream analyses.

For patients taking lipid-lowering medications, we adjusted lipid levels for medication use as described previously^39^. In particular, total cholesterol was divided by 0.8, LDL by 0.7, and triglycerides by 0.85. Due to the low frequency of individuals taking non-statin lipid-lowering medications (e.g. bile acid sequestrants), we did not adjust for these medications. In addition, we did not adjust HDL levels for medication use due to the lack of a clear quantitative relationship, although statins are generally felt to cause a modest increase in HDL levels.

### Calculation of ancestry-specific BMI cutoff values

We determined ancestry-specific BMI cutoff values with equivalent risk of type 2 diabetes as described previously^25^, except we used log transformation of BMI rather than fractional polynomials, following standard statistical practices^37^. For the outcome measure, we used the probability of type 2 diabetes generated from a logistic regression model, rather than type 2 diabetes incidence, as we were unable to reliably ascertain new diagnoses of type 2 diabetes in the biobank cohorts. We fitted a logistic regression model of type 2 diabetes status versus log(BMI), adjusted for age, sex, and genetic ancestry group. We determined the predicted probability of type 2 diabetes for an individual with European genetic ancestry and a BMI of 30. For each ancestry group, we calculated the BMI that would yield the same predicted probability of type 2 diabetes. Then, we repeated this process after adjusting the logistic regression model for the specified cluster pPS values. All tests were performed in a meta-analysis of MGB Biobank and All of Us, using a random effects model.

### Analysis of body composition metrics

For participants in MGB Biobank, we used image-based body composition metrics derived from a machine learning algorithm^40,41^. This algorithm quantifies the cross-sectional areas of muscle, subcutaneous adipose tissue (SAT), and visceral adipose tissue (VAT), as measured in abdominal computed tomography (CT) imaging at the level of the L3 vertebral body. For participants in All of Us, we used measurements of waist and hip circumference, which were measured for most participants at the time of enrollment.

## Data Availability

All referenced GWAS summary statistics are publicly available and are cited in Supplementary Tables 3 and 9. Eligible researchers can apply to access individual-level data in the All of Us program (researchallofus.org). Individual-level data in the Mass General Brigham biobank are only available with approval from the Mass General Brigham Institutional Review Board.

## Code Availability

Code for variant pre-processing, bNMF clustering, and basic visualizations is available at https://github.com/gwas-partitioning/bnmf-clustering.

## Data Availability

All referenced GWAS summary statistics are publicly available. Eligible researchers can apply to access individual-level data in the All of Us program (researchallofus.org). Individual-level data in the Mass General Brigham biobank are only available with approval from the Mass General Brigham Institutional Review Board. Code for variant pre-processing, bNMF clustering, and basic visualizations is available on the project GitHub repository.

https://github.com/gwas-partitioning/bnmf-clustering

## Acknowledgments

AJD is supported by NIH/NIDDK T32 DK007028 and NIH/NIDDK F32 DK137487. KEW is supported by NIH K01DK133637. LS is supported by funds from the Ministry of Education and Science of Poland within the project “Excellence Initiative—Research University”, the Ministry of Health of Poland within the project “Center of Artificial Intelligence in Medicine at the Medical University of Bialystok” and American Diabetes Association grant 11-22-PDFPM-03. MC is supported by the Novo Nordisk Foundation (NNF21SA0072102), and NIDDK UM1 DK126185. JMM is supported by American Diabetes Association Innovative and Clinical Translational Award 1-19-ICTS-068, American Diabetes Association grant #11-22-ICTSPM-16 and by NHGRI U01HG011723. MSU is supported by NIDDK K23DK114551, NIDDK R03DK131249, and Doris Duke Foundation Award 2022063.

We thank the Meta-Analyses of Glucose and Insulin-related traits Consortium (MAGIC) for providing pre-publication access to GWAS summary statistics for post-challenge insulin resistance measures.

## Author contributions

Conceived and designed the study: KS, AJD, MSU

Conducted analysis: KS, AJD, CM

Curated data: KS, AJD, HK, SH, RM, PHS, KEW, LS, TDM, VK, AW, AKM, JMM

Provided feedback on analysis: MC, JCF, AKM, JMM, KJG, MSU

Wrote initial draft: KS, AJD, MSU

All co-authors approved the final version of the paper

