## Extended Data Figures for "Multi-ancestry Polygenic Mechanisms of Type 2 Diabetes Elucidate Disease Processes and Clinical Heterogeneity"

**Extended Data Fig. 1. Overview of high-throughput bNMF pipeline for multi-ancestry (MA) clusters.** (A) Flowchart of the steps implemented in Part I of the pipeline, where the variant preprocessing is primarily completed. Steps include: 1) extract variants from diverse set of T2D GWAS datasets, 2) apply LD-pruning across reference panels for all populations included, to ensure independent genetic signals, 3) find proxy variants for variants that are multi-allelic, ambiguous, or have low trait counts, and 4) align variants to risk increasing alleles in largest MA T2D GWAS and remove if their *P* value in this GWAS does not meet a Bonferroni threshold. (B) Flowchart of the steps implemented in Part II of the pipeline, where the trait preprocessing is primarily completed. Steps include: 1) filter trait GWAS by minimum sample size, 2) filter trait GWAS by a minimum Bonferroni-corrected *P* value across the selected variants, 3) filter by correlation between traits and 4) generate a variant by trait association matrix.

**B.**

**A.**

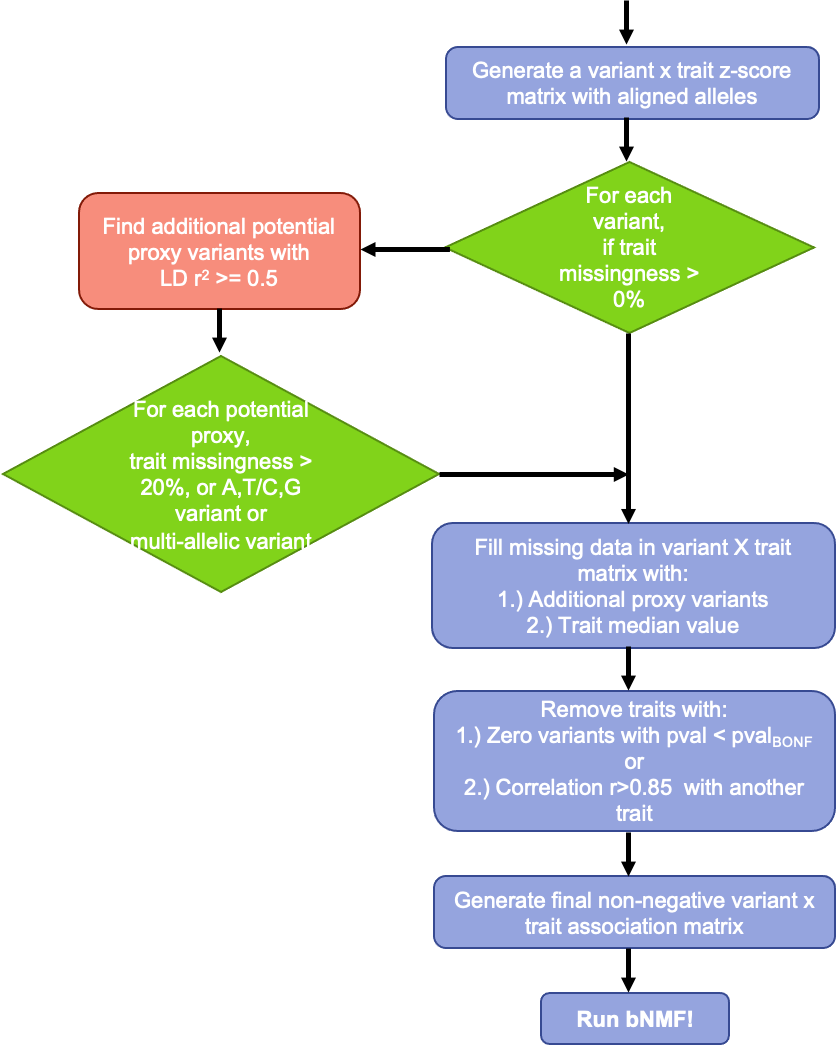

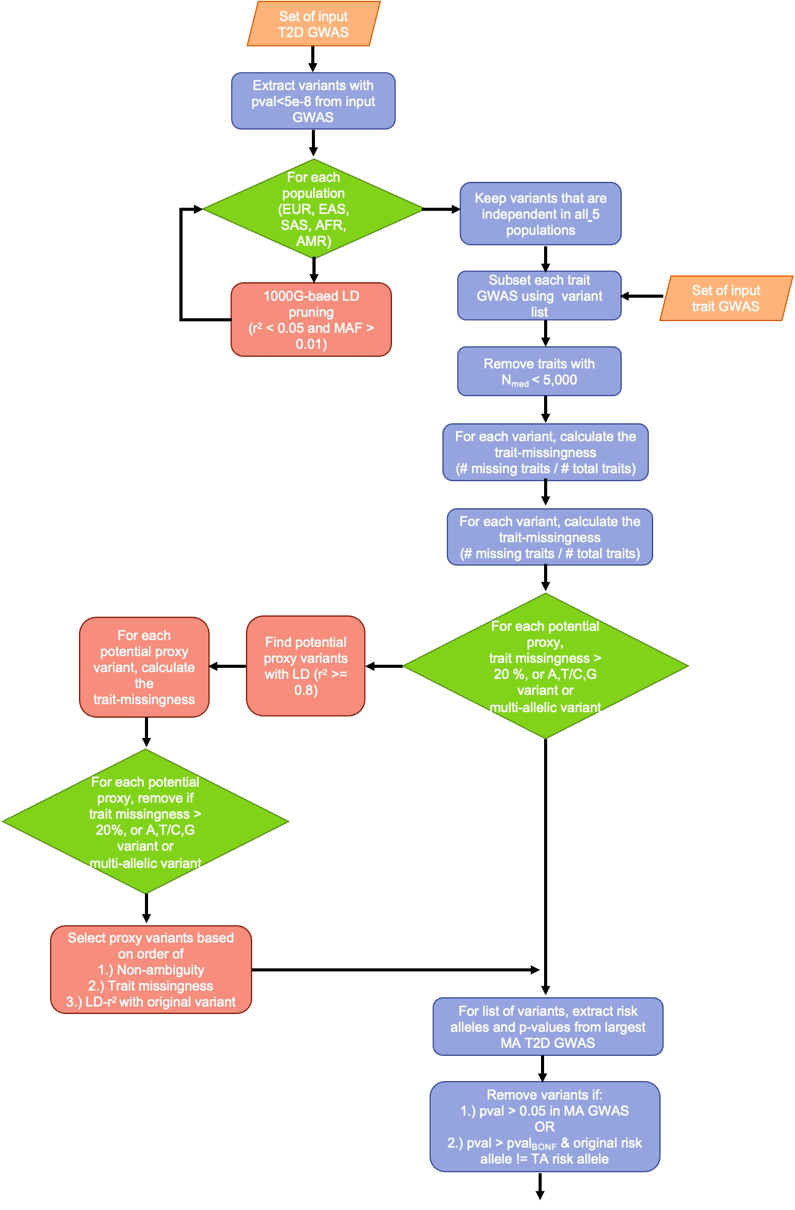

**Extended Data Fig. 2. The multi-ancestry clusters recapture several key pathways that were identified in our previous papers.** Correlation heatmap for trait cluster weights from the multi-ancestry clusters versus the (A) Udler *et al.* clusters^3^ and (B) Kim *et al.* clusters^4^. Correlation coefficients are displayed for trait pairs where *R* > 0.

**B.**

**A.**

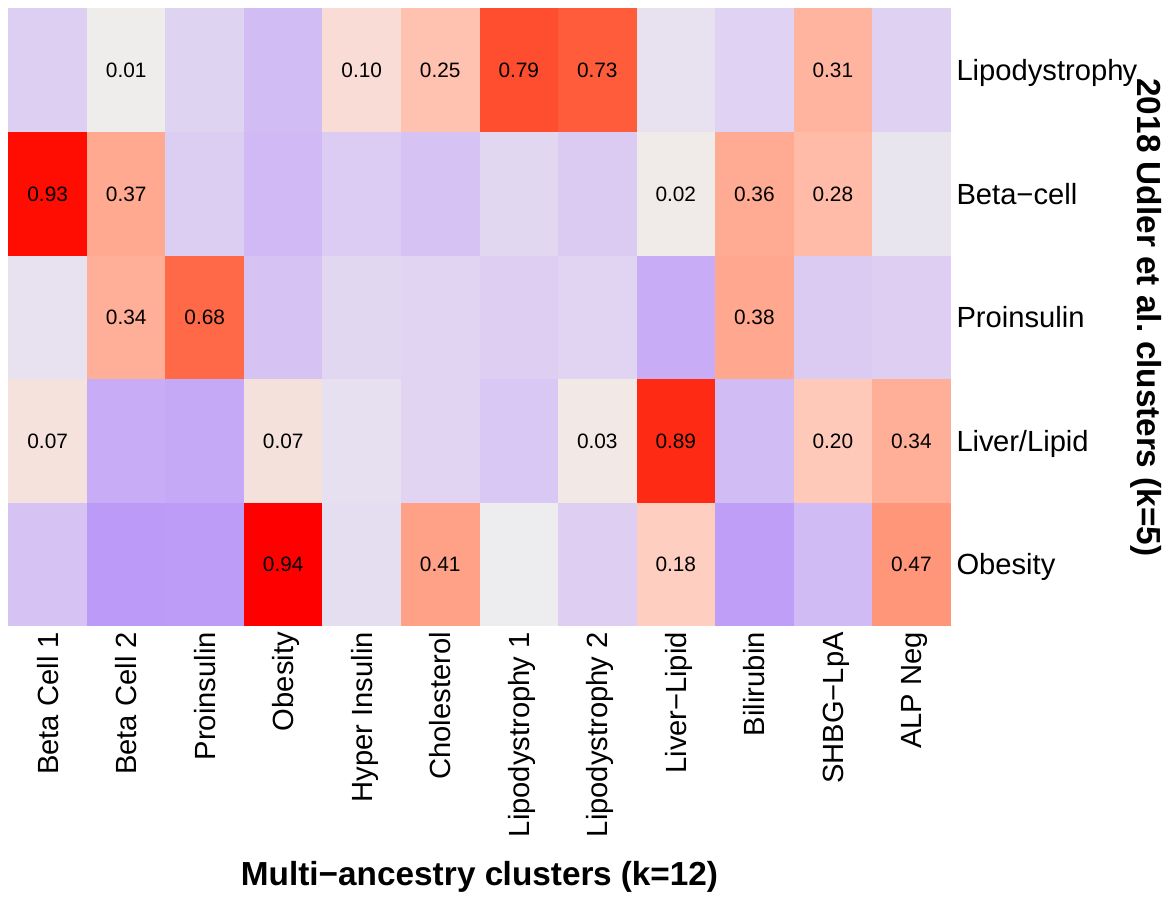

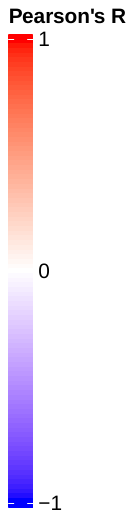

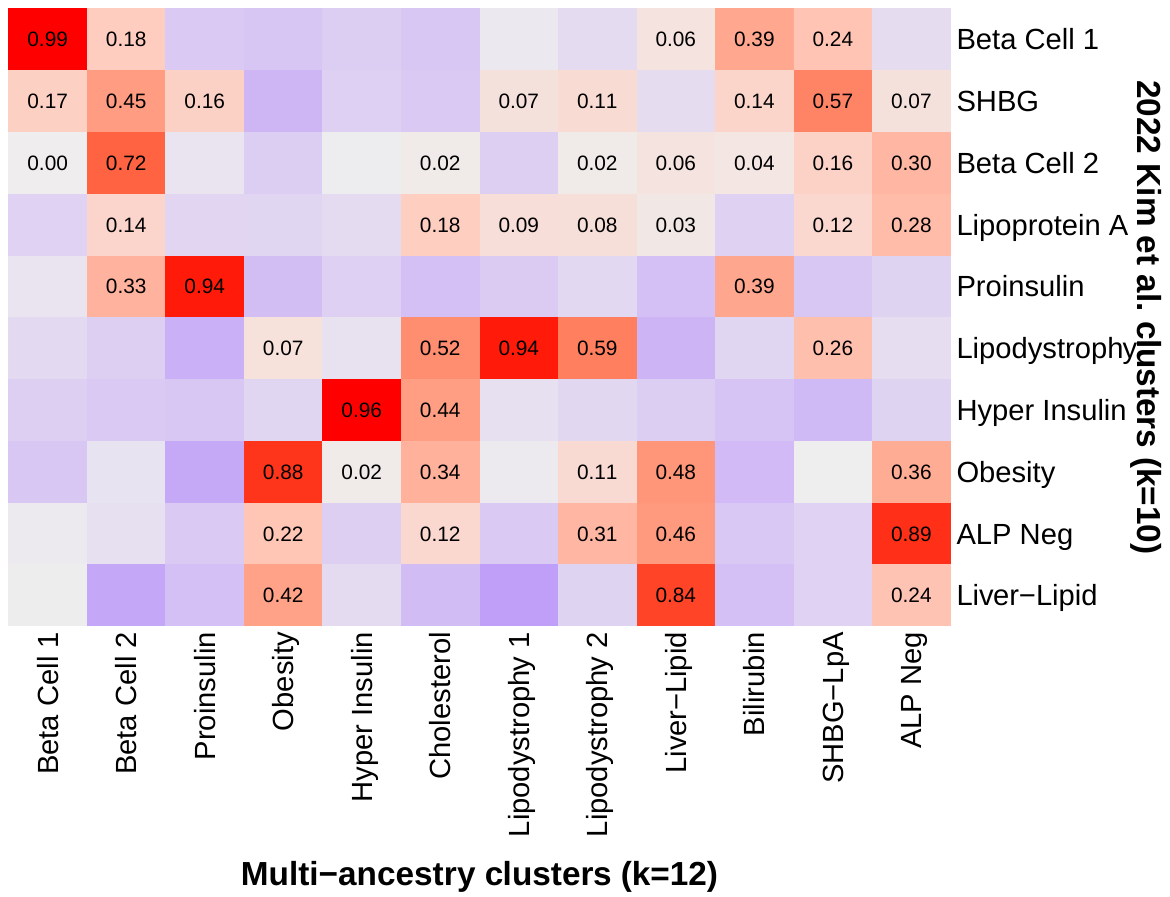

**Extended Data Fig. 3. Common T2D genetic clusters are shared across individual ancestry groups.** Heatmaps display the correlation of the trait cluster weights in the European, East Asian, African and Admixed American clusters versus the (A) multi-ancestry and (B) Udler et al. clusters^3^.

**A.**

**B.**

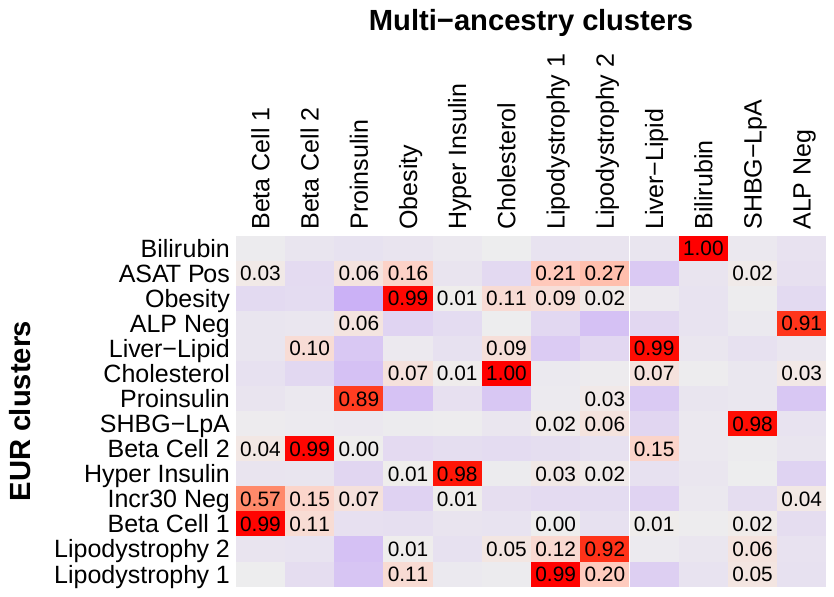

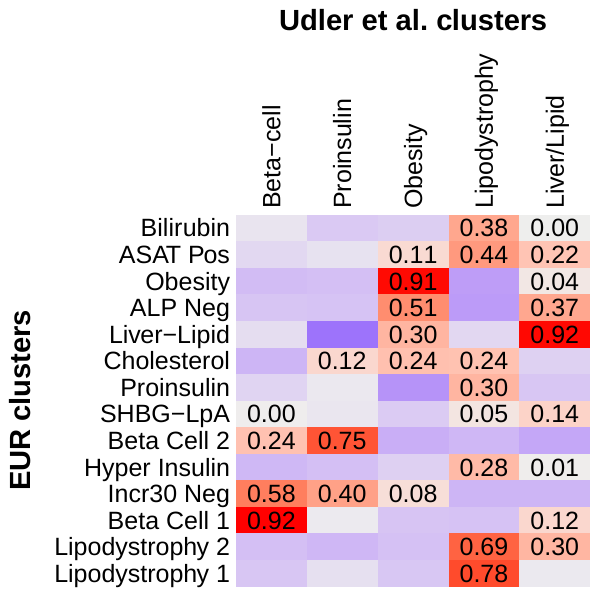

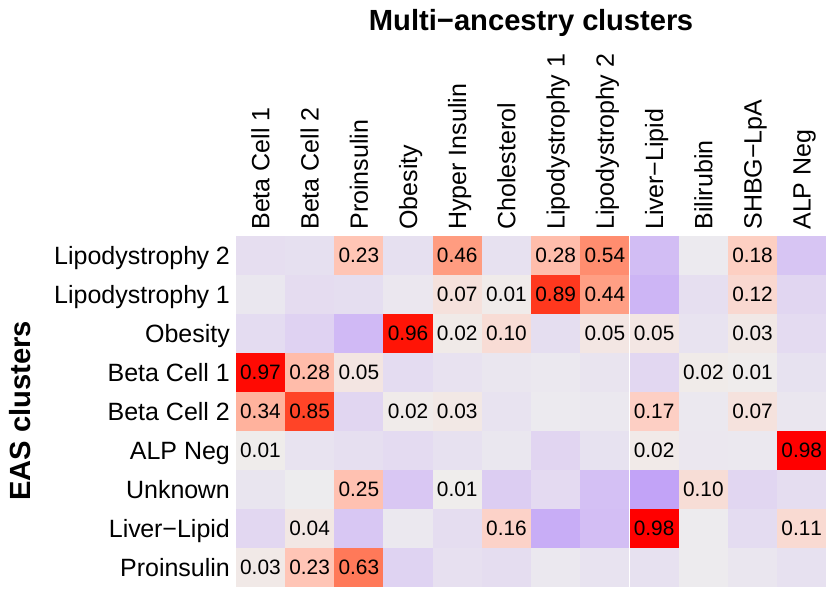

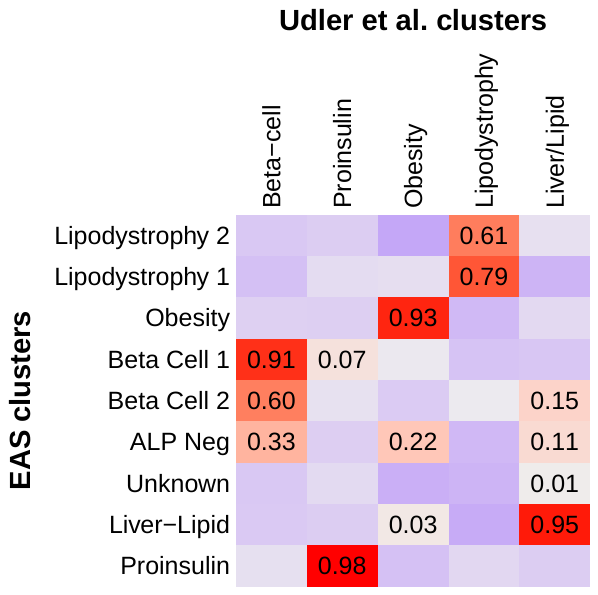

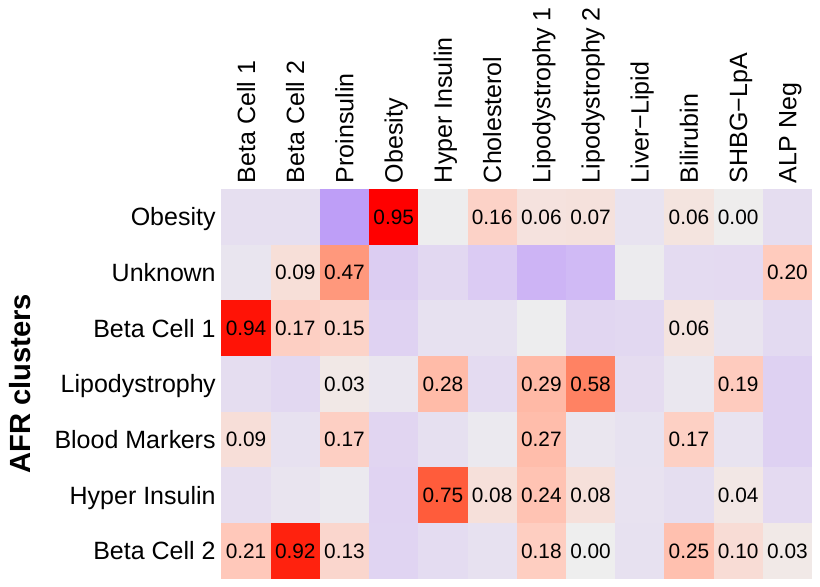

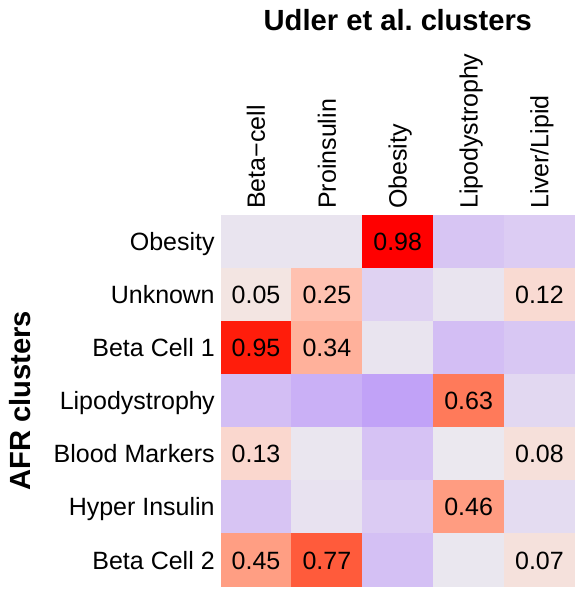

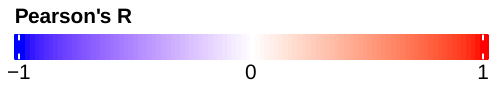

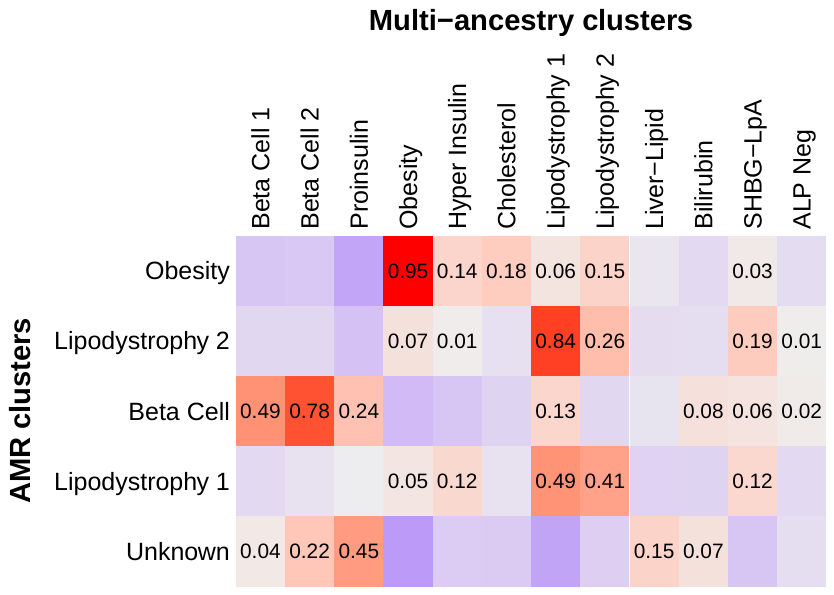

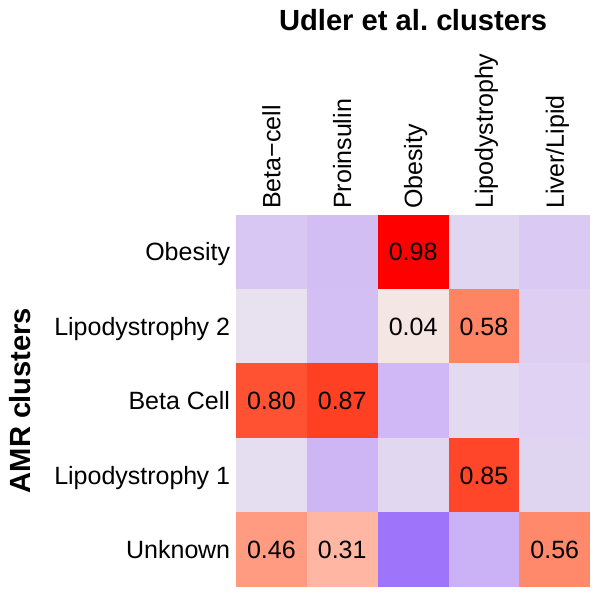

**Extended Data Fig. 4. Sex-stratified association of multi-ancestry T2D genetic clusters with visceral and subcutaneous adipose tissue.** Each bar displays the sex-stratified association between selected multi-ancestry T2D cluster pPS and selected measures of adipose distribution (visceral adipose tissue [VAT], subcutaneous adipose tissue [SAT], or VAT/SAT ratio). Each outcome was normalized to a standard normal distribution (for all participants, females only, or males only). Effect sizes indicate the effect per one standard deviation increase in the pPS. Error bars denote the standard error. Analyses were performed in a subset of approximately 9,000 MGB Biobank participants with available data on adipose distribution. * *P* < 0.05.

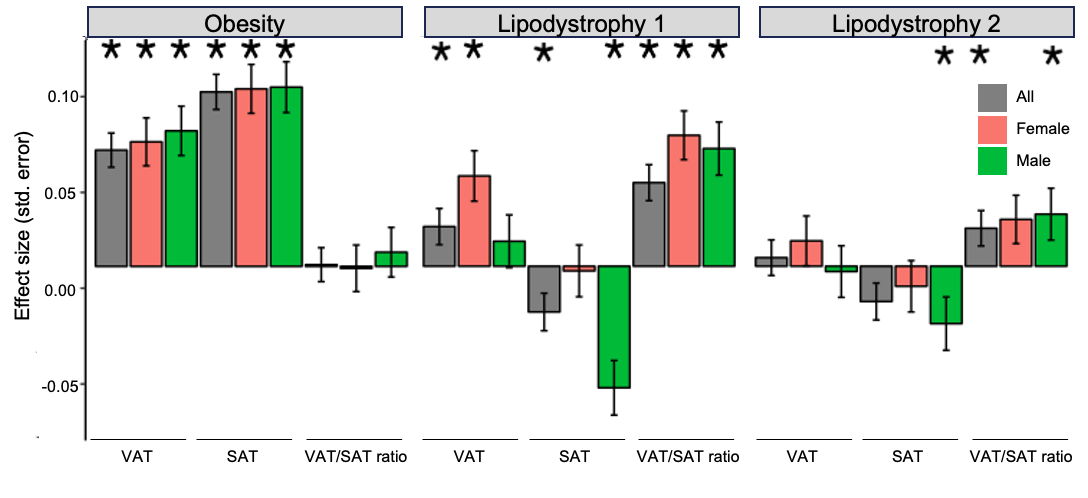

**Extended Data Fig. 5. Sex-stratified association of multi-ancestry T2D genetic clusters with waist and hip circumference.** Each bar displays the sex-stratified association between selected multi-ancestry T2D cluster pPS and selected anthropometric measures (waist circumference, hip circumference, or waist/hip ratio). Each outcome was normalized to a standard normal distribution (for all participants, females only, or males only). Effect sizes indicate the effect per one standard deviation increase in the pPS. Error bars denote the standard error. Analyses were performed in All of Us only. * *P* < 0.05.

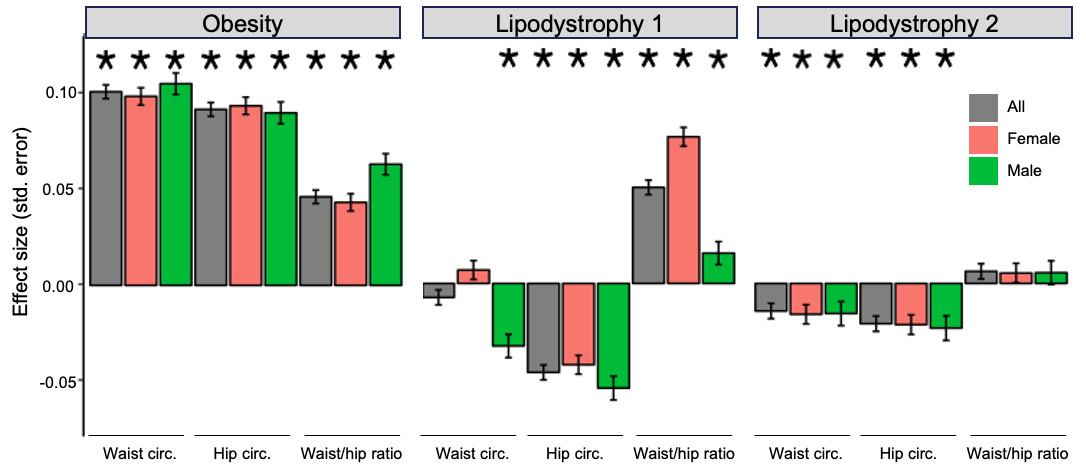

**Extended Data Fig. 6. Variation in distribution of multi-ancestry T2D genetic clusters across ancestry groups**. Each histogram displays the distribution of the pPS for the indicated multi-ancestry T2D genetic cluster. For each cluster, the pPS for the entire cohort was normalized to a standard normal distribution, and a separate curve is displayed for each genetically inferred ancestry group. All analyses were performed in a meta-analysis of All of Us and MGB Biobank. AFR, African; AMR, Admixed American; EAS, East Asian; EUR, European; SAS, South Asian.
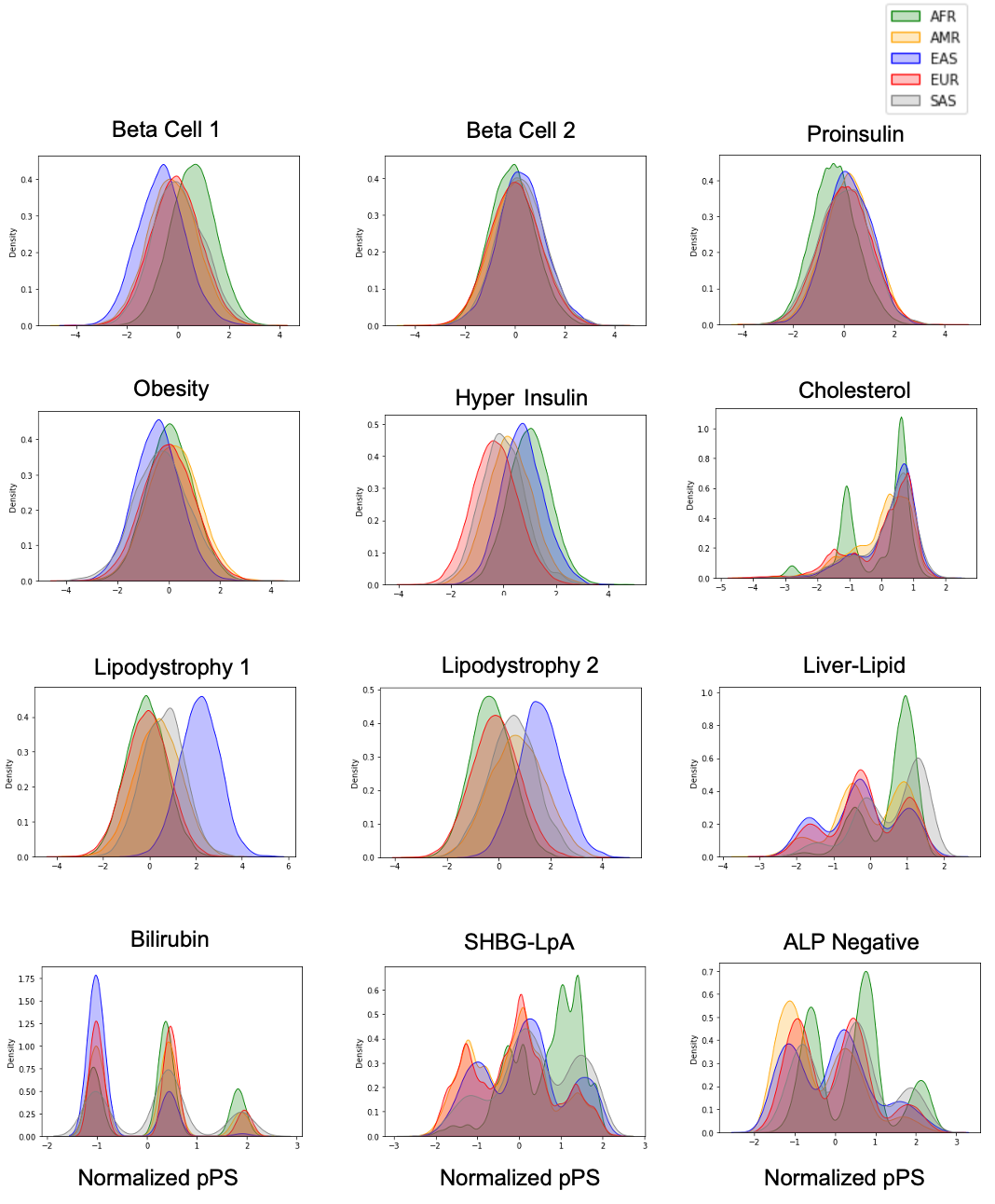

**Extended Data Fig. 7. Proportion of total T2D genetic risk attributable to each multi-ancestry T2D cluster.** For each individual, the total T2D genetic risk was calculated as the sum of the pPS across each of the 12 multi-ancestry T2D genetic clusters. All individuals were then grouped according to genetically inferred ancestry. Each bar displays the proportion of the total T2D genetic risk conferred by each specific cluster. This graph represents a meta-analysis of All of Us and MGB Biobank. AFR, African; AMR, Admixed American; EAS, East Asian; EUR, European; SAS, South Asian.

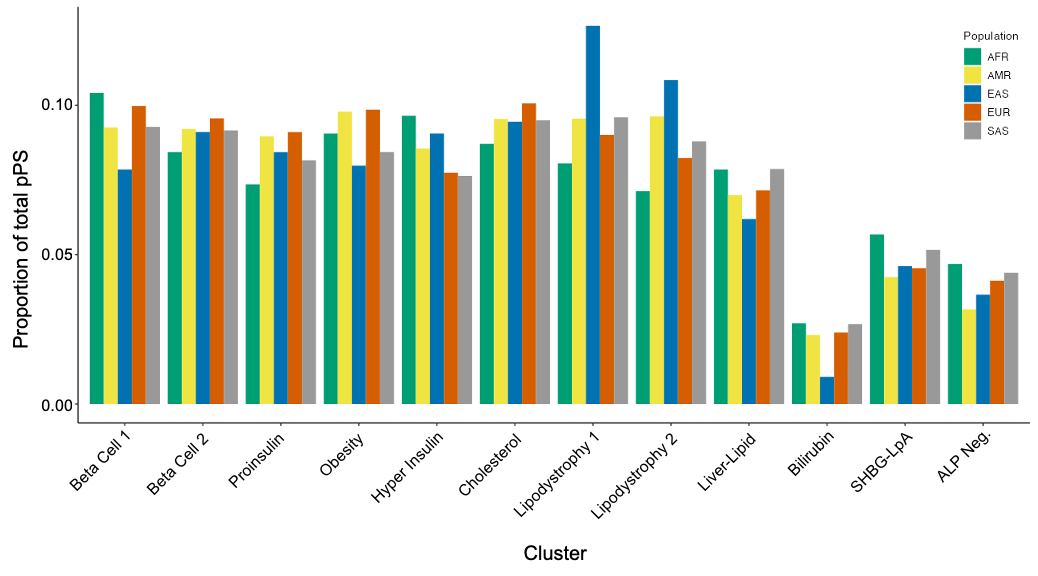

**Extended Data Fig. 8. Conservation of biological pathways between the multi-ancestry and T2DGGI clusters.** Variants in the T2DGGI clusters (*N_SNP_* = 1,289) were assigned a proxy variant from the multi-ancestry variant set (*N_SNP_* = 650) based on linkage disequilibrium values (*r*^2^ > 0.5). The clusters were then cross-examined using the Wilcoxon rank-sum test based on the multi-ancestry bNMF variant weights.

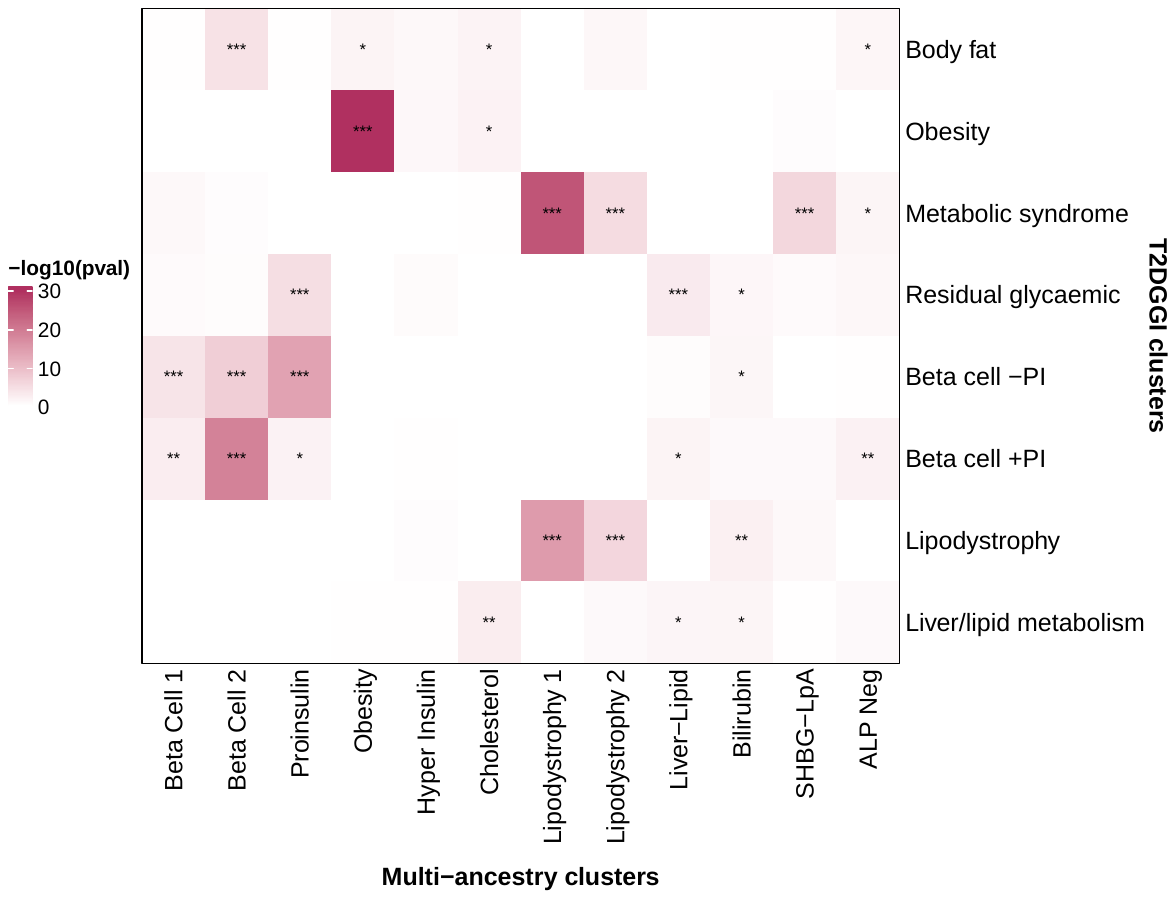

*** pval < 0.001

** pval < 0.01

* pval < 0.05
